# Leveraging global genomic sequencing data to estimate local variant dynamics

**DOI:** 10.1101/2023.01.02.23284123

**Authors:** Zachary Susswein, Kaitlyn E. Johnson, Robel Kassa, Mina Parastaran, Vivian Peng, Leo Wolansky, Samuel V. Scarpino, Ana I. Bento

## Abstract

Accurate, reliable, and timely estimates of pathogen variant risk are essential for informing public health responses. Unprecedented rates of genomic sequencing have generated new insights into variant dynamics. However, estimating the fitness advantage of a novel variant shortly after emergence, or its dynamics more generally in data-sparse settings, remains difficult. This challenge is exacerbated in countries where surveillance is limited or intermittent. To stabilize inference in these data-sparse settings, we develop a hierarchical modeling approach to estimate variant fitness advantage and prevalence by pooling data across geographic regions. We demonstrate our method by reconstructing SARS-CoV-2 BA.5 variant emergence, and assess performance using retrospective, out-of-sample validation. We show that stable and robust estimates can be obtained even when sequencing data are sparse. Finally, we discuss how this method can inform risk assessment of novel variants and provide situational awareness on circulating variants for a range of pathogens and use-cases.

## INTRODUCTION

Viral emergence, diversity, and circulation can impact outbreak dynamics and control efforts both globally and locally. In disease systems as diverse as influenza virus, HIV, and dengue virus (DENV), competition between phenotypically distinct subpopulations drives disease dynamics^1–3^. For example, population-level DENV dynamics and individual-level disease severity are both driven by the interaction between immunity and co-circulating DENV serotypes^4–6^. In the case of SARS-CoV-2, emergence of novel variants with sufficient antigenic distance from prior variants has led to surges in cases and altered vaccination strategies^7–10^. The emergence and spread of these variants have been closely tracked, sparking investigations into variant risk assessment (e.g., fitness advantage), mechanisms driving advantage (e.g., immune evasion, binding properties that enhance infectivity/transmissibility), and regional estimates of variant prevalence to inform public health. These investigations have been enabled via an unprecedented level of genome sequencing and sharing^11–13^.

SARS-CoV-2 has been sequenced in greater numbers and with greater frequency than any other pathogen^12^. The resulting information has informed public health practice^7, 14–16^, enhanced disease forecasting models^17^ and aided vaccines, therapeutics, and molecular diagnostic assay development^18, 19^. Research contributions have included detailed investigations of variant invasion and source-sink dynamics^7, 15, 16, 20^, characterizations of the contribution of specific single nucleotide polymorphisms to population-level transmission dynamics^21, 22^, and examinations of the change in relative variants’ fitness across multiple waves of transmission^23–25^. To inform mitigation and control measures and provide local situational awareness, public health agencies have produced real-time reports on emerging variants and local variant prevalence^26–30^. An understanding of variant properties (e.g., immune evasion, transmissibility, and therapeutic efficacy) combined with accurate regional estimates of each variant’s prevalence has been applied to optimize public health strategies to mitigate transmission and disease^14, 16, 21, 31–36^.

Accurately estimating the relative fitness of novel or emerging variants is a challenging problem, both for infectious diseases broadly and for SARS-CoV-2 specifically. Most genomic sequencing is performed in the Global North, limiting global breadth of genomic epidemiology for all types of pathogens^37^. In the case of SARS-CoV-2, sequencing concentrated into a few countries — 90% of publicly available SARS-CoV-2 sequences come from just 10% of countries (Figure 1). These disparities are not only in the raw number of sequences available, but also in the percent of cases sequenced and the turnaround time from sample collection to sequence availability^12^, which further limits the amount of information available for real-time or near-real time analyses. We note that this sparsity in sequencing can arise through a number of different mechanisms, including a limited local number of genomic sequences (i.e., “sequencing effort”) or limited ability to divert laboratory resources from existing use cases toward a pathogen of public health interest (i.e., “sequencing capacity”)^37^. Estimation of relative fitness based on data from only those countries with high sequencing rates delays characterization of novel variants and biases estimates toward those countries’ particular immune landscapes. Furthermore, global sequencing rates of SARS-CoV-2 have begun to fall, further obscuring and delaying our understanding of emerging variants^37, 38^.

**Figure 1.**
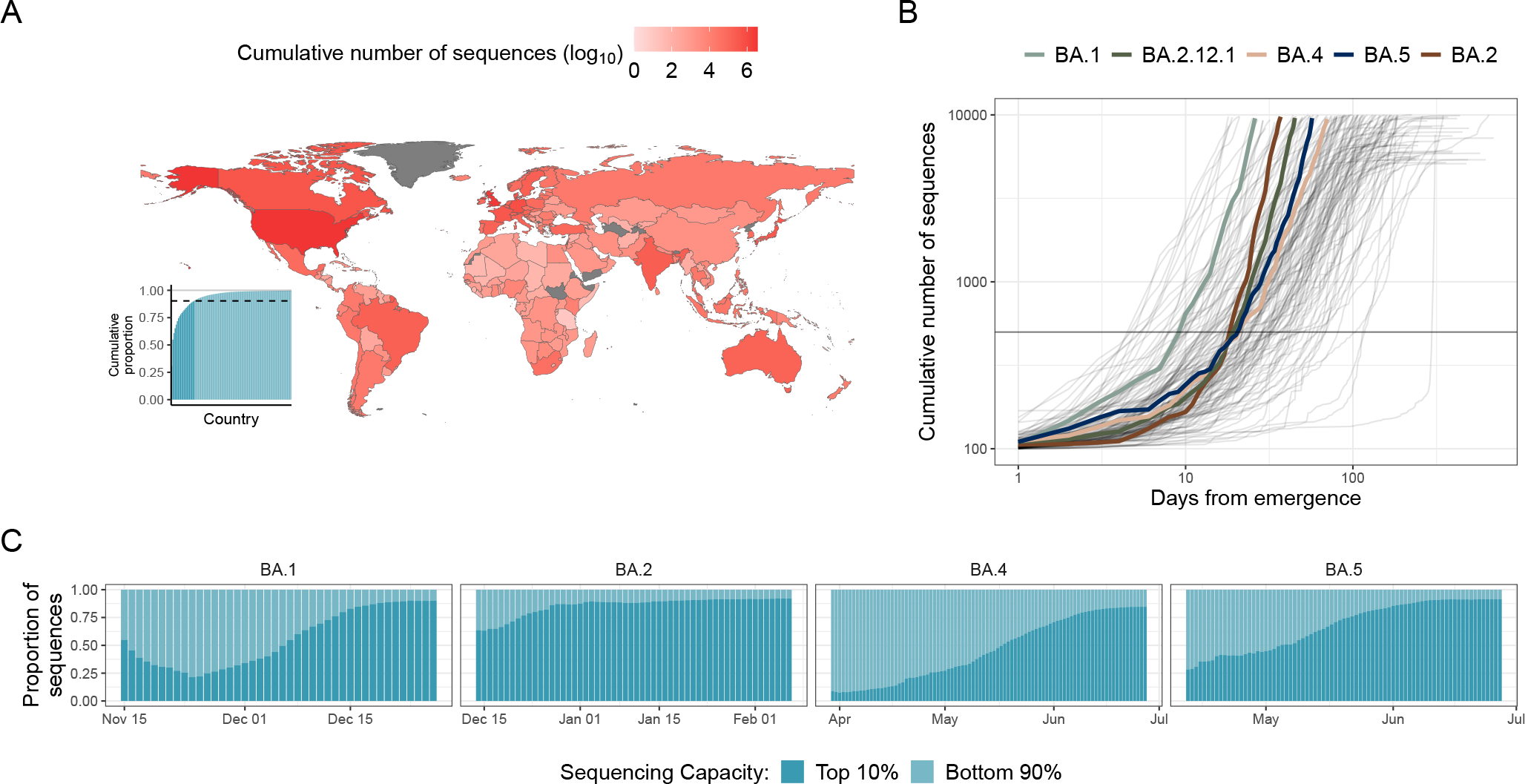
Landscape of SARS-CoV-2 genomic data and early emergent variant dynamics. (A) Shading indicates cumulative number, in log_10_ scale, of SARS-CoV-2 sequences submitted to GISAID as of July 1^st^, 2022^13^. (inset) Cumulative proportion of all sequences, with countries ordered by their relative contribution. Light blue indicates countries above the top 10^th^ percentile of contributions, dark blue indicates countries below the bottom 90^th^ percentile of countries. (B) Cumulative number of sequences versus days from variant emergence, with variants of interest which grew rapidly after emergence highlighted in color. Gray horizontal line at 500 sequences is included to highlight the time it took to reach this level for the key variants. (C) Proportion of sequences sampled by the high sequencing capacity countries (light blue) vs the lower sequencing capacity countries (dark blue) over time, starting from after a variant’s emergence.

Across disease systems, early characterization of emerging variants is constrained by data sparsity and heterogeneous diagnostic and sequencing capacity across the globe. To address this challenge, we developed a Bayesian nested hierarchical modeling approach that jointly estimates the trajectories of variants’ growth over time. From this model, we identify the key quantities of: *i*.) within-country prevalence, *ii*.) within-country relative variant fitness advantage, and *iii*.) global fitness advantage relative to circulating variants. The method accounts for the observed heterogeneity in variant fitness across geographic regions and sparsity in sequencing to provide more robust estimates of variant fitness shortly after emergence. We apply it to quantify and dissect, at a global scale, the emergence and rapid takeover of the BA.4 and BA.5 variants from the previously dominant BA.2 variant from April to July of 2022. We demonstrate our method’s ability to improve early risk assessment for novel variants and increase availability of real-time variant prevalence estimates in countries with lower sequencing capacity.

## RESULTS

### Global landscape of SARS-CoV-2 genomic surveillance

In Figure 1, we examine the landscape of global SARS-CoV-2 genomic sequencing, identifying systemic variability in sequencing capacity/throughput^13^. The majority of the world’s SARS-CoV-2 sequencing has occurred in the United States and United Kingdom, which had contributed 54.9% of all SARS-CoV-2 sequences shared via the GISAID Initiative as of July 1, 2022. This disparity extends beyond these two countries — 10% of the countries that have shared sequences with GISAID have produced 90.3% of the SARS-CoV-2 sequences (Figure 1A). These high sequencing-capacity countries are unevenly geographically distributed and generally skewed towards historically well-resourced countries; the lower-middle income country of India is notable exception. In contrast, most countries in the African continent and the Middle East have submitted only a few thousand SARS-CoV-2 sequences in total, with some notable exceptions like South Africa and Kenya (Figure 1A, inset).

In Figure 1B, we demonstrate a pattern in sequencing effort targeted towards emerging variants finding that for each variant the rate of sequencing accelerates over time. Early on, when a variant is at low prevalence, the total number of sequences of that variant grows slowly — at a linear rate. In the early stages of variant emergence, the amount of information about the variant is limited, with only a few sequences available and the benefit provided by pooling information is highest. Later on, when that variant begins to spread more widely, we see exponential growth in the total number of sequences. This pattern of an increasing rate of sequencing over time leads to the characteristic elbow shape in the plot.

In Figure 1C, we show that this second phase of rapid sequence collection is largely driven by the concentration of genomic sequencing within a few high-capacity countries. Shortly after variant emergence, the majority of the BA.1, BA.4, and BA.5 lineage sequences collected were from outside of these high sequencing capacity countries. However, once these variants reached high sequencing capacity countries, sequences were collected more rapidly, becoming a majority of the sequences collected. This change highlights that genomic surveillance for emerging variants has been largely limited to a few high-resource geographies.

In Figure 2, we describe our method for estimating multi-strain dynamics and relative fitness advantages by partially pooling information across patches (geographic subunits, e.g., countries) and strains. This statistical approach leverages a hierarchical mixed-effects Bayesian framework. The model has two levels of hierarchy. In the first level, country-specific variant fitness advantages are structured such that the fitness advantage of a variant in one location informs the expected fitness advantages of variants in other locations to formalize the assumption that zvariants’ relative fitness in one location are likely to be similar to others (Figure 2A). In the second level, variants’ mean fitness advantages, averaged over countries, consist of a shared (hierarchical) normal distribution (Figure 2B). This approach shares information across variants to formalize the ecological assumption that most variants will be similarly fit to their recent ancestors and observing extreme deviations in fitness is uncommon. This assumption leads to shrinkage on extreme fitness advantage estimates for novel or otherwise infrequently observed variants for which we might otherwise overfit to noise in the data (Figure 2C).

**Figure 2.**
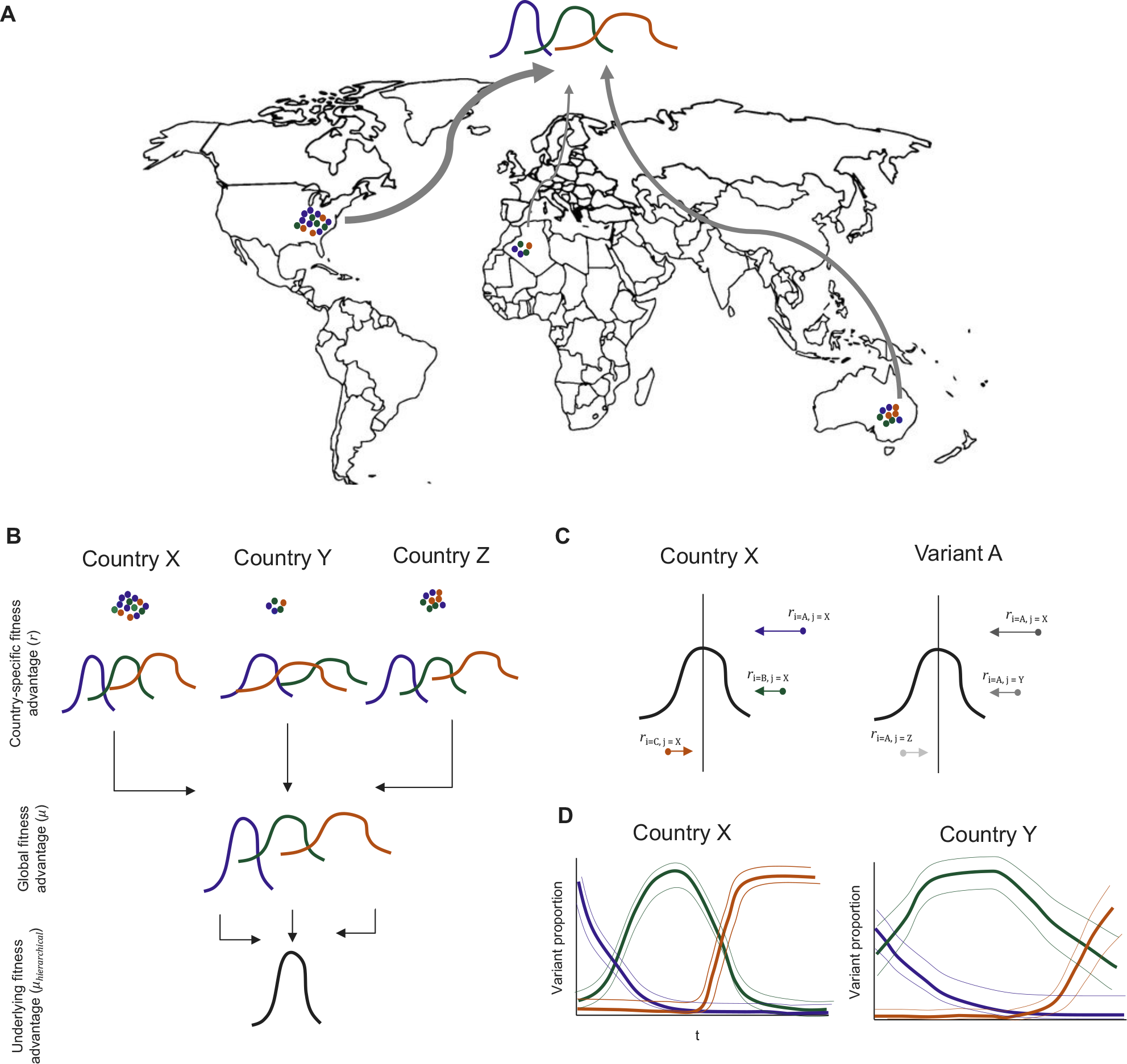
Schematic figure describing the multi-region model. (A) Observed sequences from across the globe are fit to a single Bayesian hierarchical multinomial model. (B) Model-generated posterior probability distributions of country-specific variant fitness advantages (*β*_*i j*_) are drawn from distributions of global variant fitness advantage 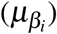 that are drawn from a global fitness distribution (*μ*_hierarchical_). Arrows indicate the hierarchical layers of the model. (C) Schematic illustration of how the mixed effects model structure results in shrinkage towards the mean of country-specific variant fitness advantages. Dots indicate estimates without pooling of information and arrows indicate the influence of hierarchical structure. (D) Model-generated country-specific estimates of variant proportions over time. Countries with fewer sequences (e.g., Country Y) will have higher uncertainty in the estimated variant proportions.

### Identifying SARS-CoV-2 variant fitness and global emergence dynamics

In Figure 3A, we present the estimated SARS-CoV-2 variant proportions alongside the number of collected sequences over time colored by the most frequently observed variant on each day for a subset of countries. Global prevalence of the BA.2 lineage declined during the period from April to July 2022, as the BA.4 and BA.5 variants rapidly took over. The invasion dynamics of BA.4 and BA.5 were heterogeneous across countries: both took over in parallel in South Africa, but BA.5 had substantially higher observed growth in Israel and Bangladesh. Notably, in Bangladesh we estimate that BA.5 had reached a higher proportion of cases (56.5% CI: 21.2-83.4%) than in India as of July 1, 2022 (13.8%, CI: 1.1-17.6%) despite their geographic proximity. In contrast, Senegal had not yet experienced substantial BA.5 invasions as of July 1, 2022. Although BA.2 prevalence declined, this change was driven by BA.4 and other lineages rather than by BA.5.

**Figure 3.**
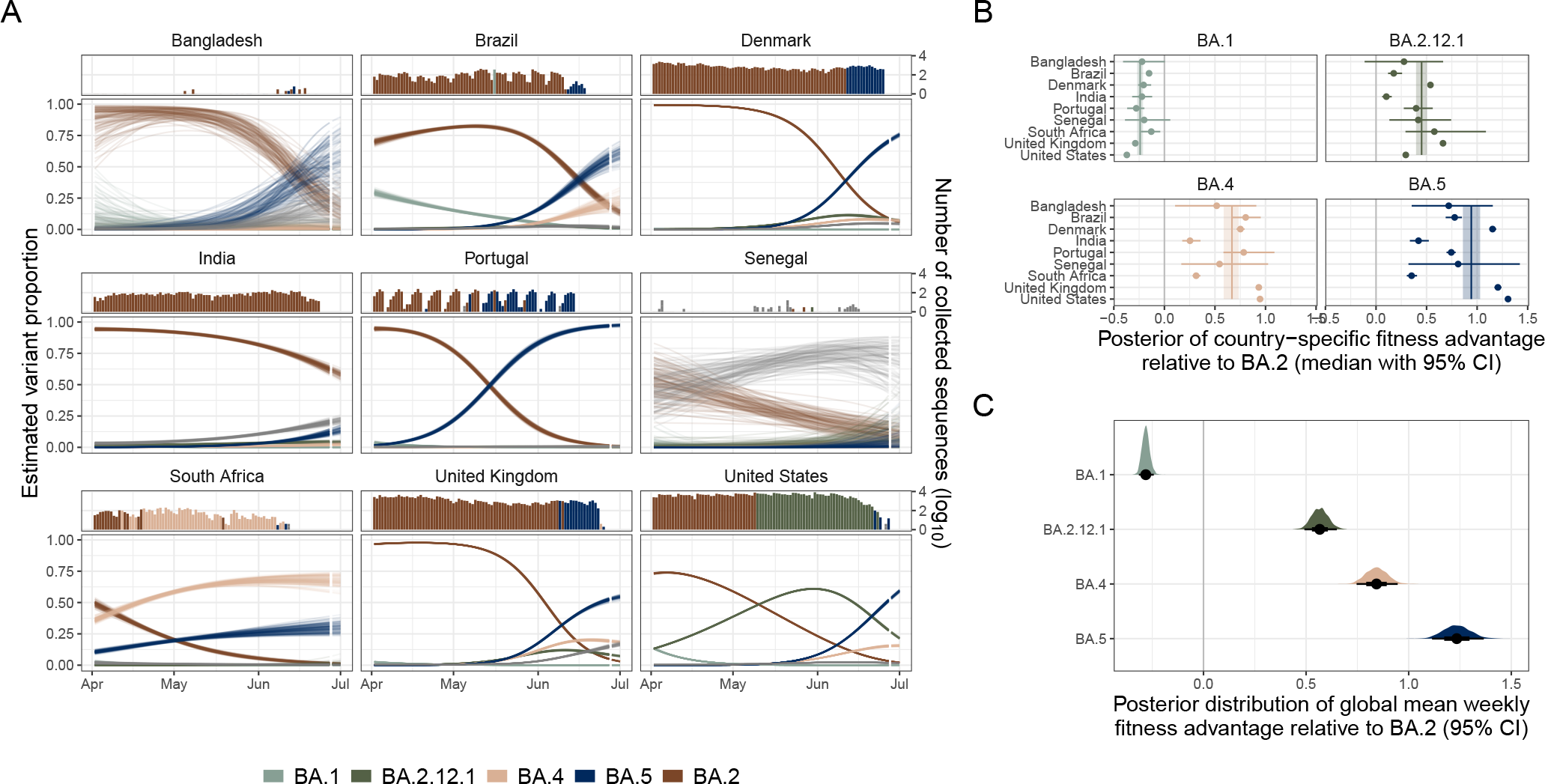
Estimating variant dynamics and fitness advantages (A) Model estimated variant dynamics in a subset of countries. Colors indicate variants, lines represent draws from the marginal posterior distributions of the country-specific estimates. The top panel shows the number of sequences collected over time, colored by the dominant variant at that time as is observed in the data. (B) Country-specific fitness advantages for selected variants (points). Vertical line indicates global estimate of variant fitness advantage. Bars and bands indicate 95% credible intervals. (C) Global posterior fitness advantage distributions for selected variants. Points indicate median, bars indicate 95% credible intervals.

In Figure 3B, we present country-specific variant growth rates, finding high country-specific fitness advantages for BA.4, BA.5, and BA.2.12.1. Country-specific fitness advantages are heterogeneous within variant type: the relative fitness of BA.4 and BA.5 are much higher in the United States (BA.5: 1.31% CI: 1.29-1.33%, BA.4: 0.95% CI: 0.92-0.97%) and the United Kingdom (BA.5: 1.21% CI: 1.17-1.24%, BA.4: 0.93% CI:0.91-0.96%) than in India (BA.5: 0.42% CI: 0.34-0.52%, BA.4: 0.25% CI: 0.18-0.36%) and South Africa (BA.5: 0.35% CI: 0.31-0.41%, BA.4: 0.25% CI: 0.18-0.36%). We note that while these estimated fitness advantages are often similar between the United States and United Kingdom (e.g., in the cases of BA.4 and BA.5), this relationship does not always hold true. In the case of BA.2.12.1, estimates of relative fitness advantage in the United States (0.29% CI: 0.29-0.30%) are much lower than in the United Kingdom (0.66% CI: 0.64-0.69%).

In Figure 3C, we present global relative fitness advantages for multiple variants of concern, finding that BA.5 is meaningfully more fit than BA.4 and BA.2.12.1. These expectations are the means of the Gaussian posterior distributions of country-specific estimates 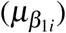 in Figure 2 (i.e., the random effect distribution) and a robust measure of overall variant fitness advantage. Most of the posterior density of the expected relative fitness advantage for BA.5 (0.94% CI: 0.85-1.03%) is higher than that for BA.4 (0.67% CI: 0.59-0.73%) which is itself higher than that for BA.2.12.1 (0.45% CI: 0.40 - 0.50%) (Figure 3C). A weekly fitness advantage of 100% means that in one week the proportion of total cases caused by BA.5 is expected to double, relative to those of BA.2 (i.e. BA.5 would go from 1% to 2% of cases if the proportion of those due to BA.2 stay constant). All three variants (BA.5, BA.4, and BA.2.12.1) have fitness advantages substantially above 0, indicating that each is more fit than BA.2, while the expected fitness advantage of BA.1 is below 0 (−0.24% CI: -0.27 to -0.21%) — indicating that it is less fit than BA.2.

### Retrospective validation and comparison

We performed a retrospective validation of the multicountry model estimates over 90 day periods using datasets from 5 reference dates. We compared the estimates from each reference date from the multicountry model to those from models fit to data from a single country (i.e., “the single country model”). Figure 4A depicts one representative comparison, using BA.5’s emergence in Portugal as a case study. We test the model’s ability to provide early, robust estimates of variant fitness using Portugal’s BA.5 emergence because Portugal had an early seeding of the BA.5 variant, prior to the variant’s widespread circulation throughout the rest of Europe. In the supplement, we show the analogous results from Figure 4A&C for additional countries (Figure S3 and Figure S4). The top panels of Figure 4A depict the sequences each model was fit to, with shading corresponding to the calibration period (i.e., the date the last available sequence from that dataset was collected). We compare the model estimates (both of the past and current variant proportions, and the 21-day forecast variant proportions) to the observed data as of July 1^st^, 2022. In Figure S1, we depict the same figure but compared to the data the model was fit to as of that reference date. This data changes over time as sequences from earlier collection dates get submitted to GISAID and “back-fill” observed variant proportions on a specific date.

**Figure 4.**
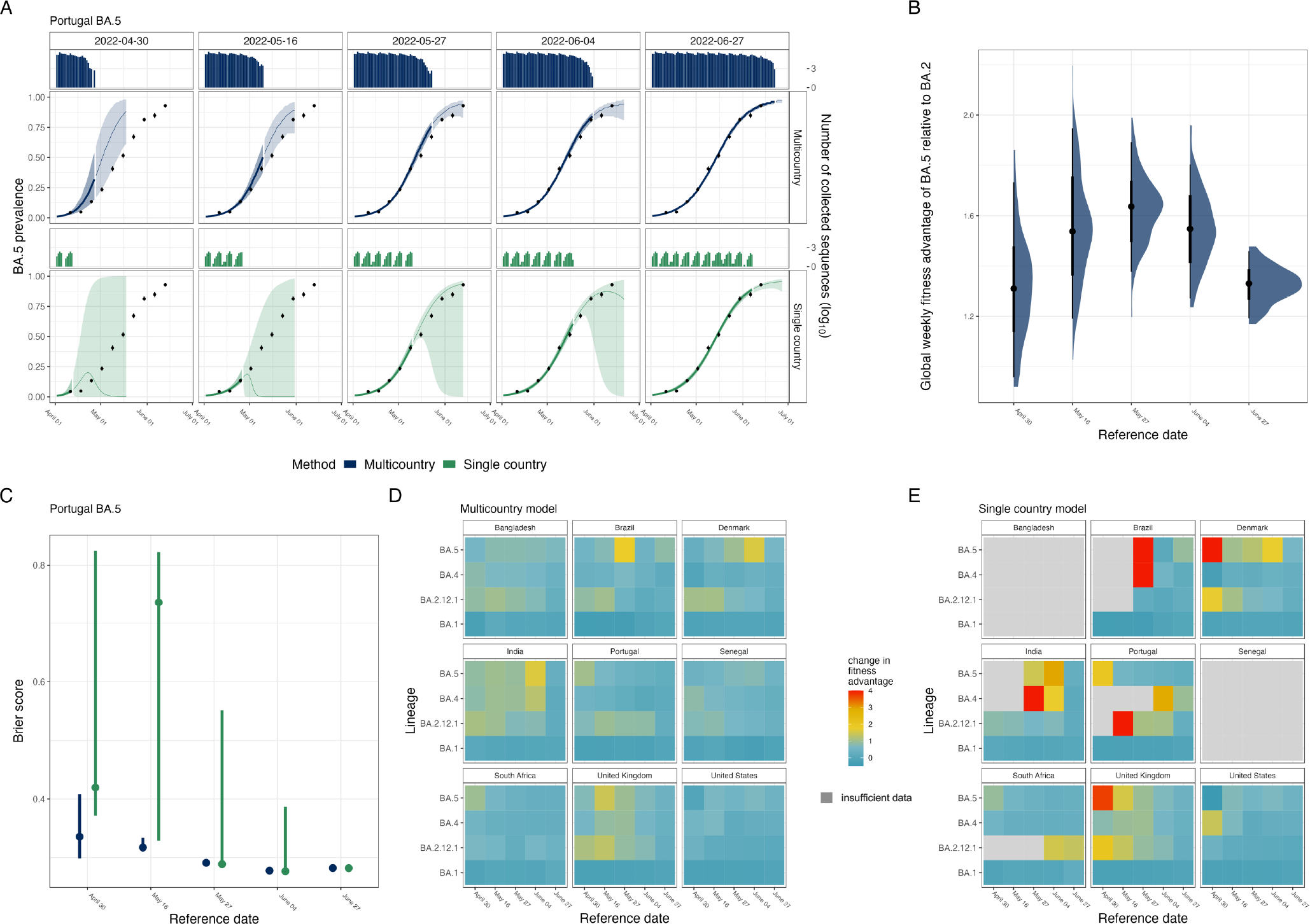
Historical validation to assess stability and predictive power of multicountry model compared to a single country model applied to BA.5 emergence in Portugal. (A) Model estimated BA.5 prevalence in Portugal from 5 successive reference dates (columns). Rows indicate whether the multicountry (blue) or single country (green) model is applied, with the bar plots indicating the number of BA.5 sequences collected by date globally or in Portugal, respectively. Uncertainty bands represent the 95% credible interval of the multicountry model and the 95^th^ percentiles of the non-parametric bootstrapped estimates for the single country model, with shading indicating the calibration period (darker) and the forecasting period (lighter). Black points indicate the observed variant prevalence as of July 1^st^, 2022, with error bars indicating *±*1 standard deviation. (B) Posterior distribution of the global estimate of BA.5’s weekly fitness advantage compared to BA.2 over time. Points indicate median weekly fitness advantage, bands indicate the 66% and 95% credible intervals. (C) Brier score evaluating predictive accuracy of estimated BA.5 prevalence to the observed prevalence on July 1^st^, 2022. Points indicate the mean Brier score, bars indicate the 95% credible interval on the Brier score from the multi-country model, and the 95^th^ percentiles of the non-parametric bootstrapped estimates for the single country model. (D)-(E). Country and variant specific changes in estimated fitness advantage from the July 1^st^, 2022 dataset compared to the estimates fit to the reference data set, for the multi-country model (D) and the single country model (E). Gray areas indicate that an estimate of the variant fitness advantage could not be made for that variant-country on that date due to insufficient data.

In Figure 4B, we show the posterior distributions of estimated global weekly mean fitness advantage of BA.5 relative to BA.2 over time (across reference dates), demonstrating that it is stable. To quantify the degree of similarity between the posterior distributions of the global weekly fitness advantage over time, we computed the Jensen-Shannon (JS) divergence comparing the 5 distributions at each reference date to the distribution from the final data (Figure S2). The maximum JS divergence was between the estimate on April 30^th^ and the estimate on May 16^th^, 2022, with a JS divergence of 0.0069. A JS divergence of 0 means the distributions are identical, and the maximum JS divergence is 1. Figure S2 depicts how the multicountry fitness advantage estimates of BA.5 in Portugal compare to the single country model fitness advantage estimates over time. The single country model estimates experience wider uncertainty at early reference dates and a sharper decline.

In Figure 4C, we evaluate the model predicted variant proportions from each reference date using the Brier score. Lower Brier scores indicate a more accurate probabilistic prediction. We compared the model predicted BA.5 proportion in Portugal to the observed proportions from the data as of July 1^st^ 2022, using the Brier score over the combined calibration and forecast periods (see the Supplementary Methods for additional detail). The multicountry model exhibits lower Brier scores for the early reference dates during variant emergence, with scores of 0.34 [95% CI:0.30-0.31] and 0.32 [95% CI:0.31-0.33] as of April 30^th^, 2022 and May 16^th^, 2022 respectively compared to scores of 0.42 [95% CI:0.37-0.82] and 0.73 [95% CI:0.74-0.82] for the single country model on those same dates.

In Figure 4D&E, we assess the stability of estimates from both the multicountry model (D) and the single country model (E) by showing the difference between the country-specific variant fitness estimate at each reference date compared to the country-specific variant fitness estimated at the final date (July 1^st^) for the two models. However, because the single country model cannot estimate country-specific variant fitness advantage if the particular variant has not been observed in sufficient quantities (<3 sequences of a variant) in that country, there are gaps in the single country model estimates indicated by the gray boxes in Figure 4E. In Figure S7, we report the number of sequences of a variant observed in each country by each reference date for context. In contrast to the gaps in the single country model (E), the multicountry model (D) is able to provide reasonable country-specific variant fitness advantage estimates even before the variant has been observed in each country. Figure S3 provides the absolute transmission advantage estimates for each country-variant combination for both models. We also evaluate a flexible, spline-based modeling approach in (Section S2.4), finding that this approach has comparable predictive performance but caution against its use to estimate relative growth rates.

The estimated country-specific variant fitness advantages from the multicountry model are more stable than those produced from the single country model (Figures S5, S6, S8, and S9). The estimated coefficients from the single country model are initially higher than those from the multicountry model and they decline to a stable estimate later than those of the multicountry model (Figure S8). Although lower, the estimated country-specific fitness advantages from the multicountry model also have an initial transient positive bias, as can be seen in Figure 4D and Figures S9 and S10. However, the estimated global mean fitness advantages are quite stable over time (Figure S10), likely due to the additional shrinkage on these estimates from the partial pooling across global variant fitness (see the Supplementary Methods for additional information).

## DISCUSSION

Enabling broader access to information from genomic surveillance for emerging and re-emerging pathogens has been and will remain a global health priority^12, 17, 19, 39–42^. In addition to the crucial work of building global sequencing capacity, methodological advances in how sequences are analyzed can address gaps in the surveillance landscape^12^. We demonstrate that our method can reliably estimate the growth of sparsely sampled competing viral variants over a short period in real time, generating both global and local estimates of variant fitness advantage and local estimates of variant prevalence. We apply this method to emerging SARS-CoV-2 variants (Figures 3 and 4, and Figures S2-S6), highlighting its robustness both for emerging variants with few sequences available and for regions with limited amounts of sequencing. We also demonstrate its applicability across a number of settings, including influenza strain dynamics to show suitability across pathogens (Figure S19) and at the state and national level in Brazil and Argentina to demonstrate relevance to local public health surveillance (Figure S20).

Academic and public health groups have leveraged the unprecedented genomic sequencing for SARS-CoV-2 to advance methods in genomic epidemiology. These advances include linking epidemiological data with genomic sequencing data to generate estimates of variant-specific *R*_*t*_^24^ or projections of the impact of specific amino acid mutations on empirical fitness dynamics using hierarchical structure — similar in spirit to the approach described here, but focused instead on deriving mechanistic insight^21, 22^. Others have applied flexible regression approaches over longer timescales or multiple variant-driven waves of incidence, reconstructing the changing fitness dynamics over time^7, 25, 43–45^. These largely retrospective analyses dissect the epidemiological drivers of variant transmission such as changes in innate transmissibility over time^25^, influence of population immunity to previous variants on secondary attack rate^7^, and relative contribution of each variant to surges in incidence^24^. These innovative analyses are mostly focus on regions with extraordinarily high-resolution genomic sampling to examine differences in variant transmission at fine spatiotemporal scales.

Real-time characterization of the growth of emerging variants and accurate nowcasting of prevalence remain challenging problems, especially in settings with sparse genomic sampling. There are often long lag times from sequence collection to submission and reporting, especially in locations with limited sequencing capacity^12^. Emerging variants by definition have few sequences available and, usually, have been detected in only a few countries. In addition, SARS-CoV-2 fitness advantage is often inflated relative to the true long-term advantage shortly after emergence, as we observe here (Figure 4, Figure S6) and has been observed empirically in other settings^46^. While the mechanisms driving this transient bias are not fully understood, we speculate that it might be driven by a combination of biases in the observation process (e.g., prioritization of samples for sequencing), stochasticity in the transmission process (e.g., superspreading early on after introduction), as well as overfitting to noisy data^47–49^. However, these mechanisms are unlikely to completely explain the consistency in the direction of the bias across the observed lineages and settings (Figures S6, S8, and S9). While beyond the scope of our method, we hypothesize that additional mechanisms may further drive these observed patterns such as network effects like crowding^50^ and preferential infection in highly social subgroups^51^.

Estimating the global risk posed by an emerging variant is a necessary step in coordinated public health responses. Accurate, real-time estimates of an emerging variant’s relative transmission advantage can focus effort for mechanistic and lab-driven studies (see e.g.,^52, 53^) as well as public health messaging. Although a variant’s local transmission is driven by the landscape of contact and immunity^44, 45, 54^, it spreads globally by an interconnected web of travel^55–57^. Effective risk ascertainment must account for these local heterogeneities in order to extrapolate to both the global and local risk posed by an emerging variant. Although a global mean weekly fitness advantage is not necessarily predictive precisely of any one locale’s variant dynamics, it provides a measure of the overall risk posed by a novel variant and can drive research and public health practice.

Although we show that our modeling approach is more robust than standard approaches (Figures S8, S9, and S11), it relies upon a number of assumptions. We assume that sequences are randomly sampled from each geographic unit of cases (e.g., country), that samples of one variant are not prioritized for sequencing over those of another variant (e.g., due to S-gene target failure in PCR testing), and that sequences are perfectly assigned to Pango lineages^58^. Further, we assume homogeneous mixing of infected individuals within a patch (e.g., country), that the relative fitness of one variant to another is linear over time on the scale of the linear predictor, and that the generation interval of each variant is the same. We note, however, that estimates of the fitness advantage can be robust to biases in sampling or sub-patch structure in disease dynamics provided that the sequences come from a consistent subset of the population. In addition, our approach of pooling data hierarchically mitigates the effects of local variability in super-spreading. These estimated fitness advantages are likely also robust to misspecification of the generation interval distribution family^20^, but the generation interval may not be constant across variants. The idealization of a constant generation interval across variants can be inaccurate^59, 60^ and can thus bias estimates of transmission advantage^61^. Further, we don’t aim to explicitly identify the very first introductions of any given variant. For a full list of assumptions and their potential implications, see the Model Assumptions and Limitations subsection of the Supplemental Methods.

Our method complements investments in genomic sequence capacity-building by better leveraging data across regions, incentivizing data sharing and providing more equitable access to real-time localized variant dynamics for pathogens with circulating strains. The global variant dynamics we uncover here can improve our understanding of variant emergence and growth and are critical for situational awareness, evidence-based decision-making, and policy design for mitigating pathogen spread.

## METHODS

### Data processing

Line list SARS-CoV-2 sequence metadata was accessed via the GISAID EpiCov database^13^. The findings of this study are based on metadata associated with 2,032,779 sequences available on GISAID up to July 1^st^, 2022, via https://doi.org/10.55876/gis8.230118ka. Using the Pango lineage annotations in the sequence metadata, we aggregated the observed sequences by country, collection date, and Pango lineage to get daily counts of the number of sequences of each lineage observed. We truncated lineage assignments to their root assignment (e.g., BA.5.1 to BA.5), but preserved specified lineages of interest (BA.1, BA.2, BA.2.12.1, BA.4, and BA.5). Lineages with fewer than 50 observed sequences in the time period of interest were marked as “Other”. Additional details on data processing are available in the Supplemental Methods.

### Hierarchical generalized linear modeling approach

We modeled the dynamics of competing variants of a directly transmitted infectious disease using a hierarchical multinomial regression approach (Figure 2). This approach can be used generally for competing strains, but here we used the example of SARS-CoV-2 variants using sequence metadata from GISAID to produce estimates of relative variant growth rates and true proportion of total cases in a given country.

We modeled the observed sequence counts *Y*_*i jt*_ of variant *i* in country *j* on day *t* as drawn from a multinomial distribution (Eq.1), with the probability *p*_*i jt*_ of observing a given variant in a country on a given day defined as the softmax of the linear predictor and *N*_*jt*_ = ∑_*i*_ *Y*_*i jt*_ (Eq. 2).

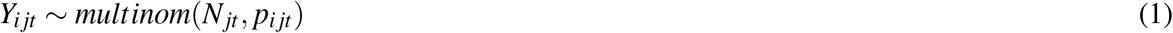

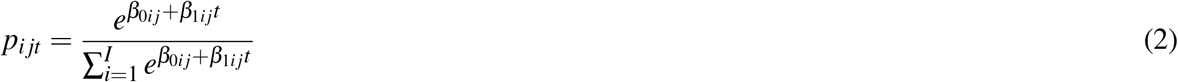

The intercepts of the linear predictor (*β*_0*i j*_) are defined as random effects drawn from a normal distribution. The intercepts describe the initial variant prevalence on the scale of the linear predictor on day 0. The relative variant growth rates for country *j* are defined as a vector of random effects drawn from a multivariate normal distribution (Eq. 3).

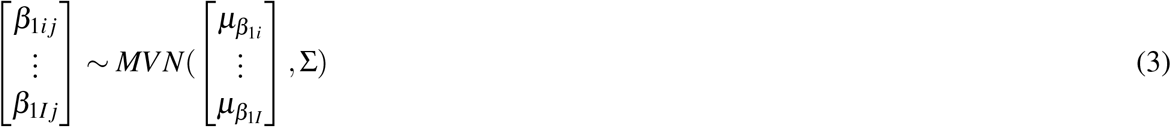

The country-specific relative growth rates *β*_1*i j*_ describe the difference in intrinsic growth rates for the variant of interest and the reference variant (i.e., where *β*_*i j*_ = *ρ*_*i j*_ *−ρ*_*I j*_, where *ρ*_*I j*_ is the intrinsic/Malthusian growth rate of the dominant variant *I* in country *j*).

The elements of the vector of the mean relative variant growth rates 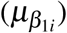 in the multivariate normal distribution are themselves random effects, drawn from a normal distribution with mean *μ*_*hierarchical*_ (Eq. 4).

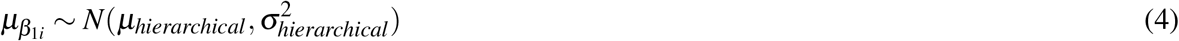

The hierarchical structure and its implications are described schematically in Figure 2. For interpretability, we presented the global and country-specific estimates of the relative growth rates as relative weekly fitness advantages using the relation as described by Davies et alia.^32^

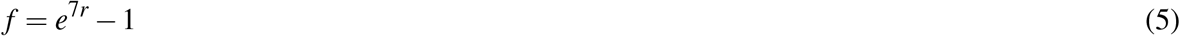

Where *r* is the relative growth rate (either *β*_1*i j*_ or 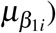) and *f* is the weekly fitness advantage displayed in the results.

This modeling approach makes a number of assumptions about the data generating process. These included random sampling of sequences within countries, that variants are correctly assigned to lineages, and that sequence collection dates are correctly reported. The modeling approach also assumed that relative variant fitness does not change over the modeled time period, such as could potentially occur due to a vaccination campaign changing the immune landscape of the host population (i.e., during the 90-day window).

Additional detail on the observation process model, model structure, prior specification, and model limitations is available in the Supplemental Methods.

### Retrospective validation of country-specific variant prevalence projections

#### Processing and fitting of historical datasets

SARS-CoV-2 line-list sequence data from GISAID was accessed on the following dates, which we refer to as “reference” dates: April 30^th^, 2022, May 16^th^, 2022, May 27^th^, 2022, June 4^th^, 2022, June 27^th^, 2022 and July 1^st^, 2022. A consensus set of lineages was found by identifying all lineages that exceed the global threshold of 50 or more observed sequences in the past 90 days across any of the 6 reference datasets. The multicountry models were fit on all countries jointly for each reference date and so used all sequences in the reference dataset. The single-country models were fit on each country separately for each reference date and so used only those sequences reported from a country in the reference datasets. For the dataset from each reference date, we defined a calibration period: the time period up to and including the day the last sequence was collected. We also defined an associated forecast period: the days after the last sequence was collected. For example, if we accessed the data on April 30^th^, 2022, and the most recent collection date in that dataset was April 27^th^, 2022, than the calibration period would be any time before and including April 27^th^, and the forecast period would be any time after April 27^th^. To test the model’s stability and predictive power, we forecast a maximum of 21 days out and compared to any data observed by the last reference dataset from July 1^st^, 2022.

#### Model comparison

For each reference dataset, we compared variant prevalence estimates from the multicountry model with estimates from the single country multinomial model. In order to estimate uncertainty, we used non-parametric bootstrapping with replacement, generating 100 bootstrapped datasets with time points randomly sampled with replacement from the true data. To fit the single country model to the data and the boot-strapped datasets, we used the nnet package v7.3-17^62^ in R v4.2.1 which returns the maximum likelihood estimation (MLE) of the multinomial model parameters, which we transform into variant fitness advantages and variant prevalence dynamics using equations 2 and 5.

#### Evaluation of model estimates

For both the multicountry and the single country model, we used 100 draws from the distributions of lineage prevalence estimates to evaluate the accuracy of the model predictions compared to the observed daily lineage prevalence from the July 1^st^, 2022 reference dataset. For countries that observed no sequences of a particular lineage in the consensus dataset during the 90-day time window, the single country estimation model does not estimate a prevalence for that lineage. To enable a fair comparison of the two model outputs at the country level, we collapsed all prevalence estimations from the multicountry model for these unobserved lineages into “other” for that country. We used the Brier score to evaluate the accuracy of each draw of the model output. The Brier score was calculated at the country-level over the combined calibration and forecast periods. The result was a distribution of Brier scores at each reference time point.

## Data Availability

All code is made publicly available at this Github repository: https://github.com/PandemicPreventionInstitute/ppi-variant-tracker-manuscript. We do not include any data in the repository, but all results can be reproduced using the GISAID SARS-CoV-2 and flu metadata for authenticated users.

## ACKNOWLEDGEMENTS

We gratefully acknowledge all data contributors, i.e., the Authors and their Originating laboratories responsible for obtaining the specimens, and their Submitting laboratories for generating the genetic sequence and metadata and sharing via the GISAID Initiative^13, 63, 64^, on which this research is based. We acknowledge the financial support of The Rockefeller Foundation which funded this work. We thank Nick Reich, Spencer Fox, and Moritz Kraemer for their valuable comments. We also gratefully acknowledge Our World in Data^65^, who curated data enabling this work.

## Author contributions statement

ZS, KEJ, SVS, and AIB designed the study, ZS, KEJ, SVS, LW, and AIB conceptualized the work, ZS and KEJ wrote the code, conducted the analyses and wrote the first draft. ZS, KEJ, SVS, and AIB contributed to the model formulations and interpreted the results. All authors contributed to editing the manuscript and gave final approval for publication and agree to be held accountable for the work performed therein.

## Notes

### Competing Interest Statement

The authors have declared no competing interest.

### Funding Statement

We acknowledge the financial support of The Rockefeller Foundation, who funded this work.

### Summary of Updates

The manuscript has been revised to: - Include additional reference to the literature. - Clarify the situations in which the described method is most appropriately applied. - Add additional validation and comparison to existing methods.

## References

1. Earn, D. J. D., Dushoff, J. & Levin, S. A. Ecology and evolution of the flu. Trends Ecolx. Evol. 17, 334–340 (2002).

2. Taylor, B. S., Sobieszczyk, M. E., McCutchan, F. E. & Hammer, S. M. The challenge of HIV-1 subtype diversity. N. Engl. J. Med. 358, 1590–1602 (2008).

3. Andraud, M., Hens, N., Marais, C. & Beutels, P. Dynamic epidemiological models for dengue transmission: A systematic review of structural approaches. PLoS One 7, e49085 (2012).

4. Recker, M. et al. Immunological serotype interactions and their effect on the epidemiological pattern of dengue. Proc. Royal Soc. B: Biol. Sci. 276, 2541–2548 (2009).

5. Katzelnick, L. C. et al. Antibody-dependent enhancement of severe dengue disease in humans. Science 358, 929–932 (2017).

6. Adams, B. et al. Cross-protective immunity can account for the alternating epidemic pattern of dengue virus serotypes circulating in bangkok. Proc. national academy sciences 103, 14234–14239 (2006).

7. Kraemer, M. U. G. et al. Spatiotemporal invasion dynamics of SARS-CoV-2 lineage b.1.1.7 emergence. Science 373, 889–895 (2021).

8. DeGrace, M. M. et al. Defining the risk of SARS-CoV-2 variants on immune protection. Nature 605, 640–652 (2022).

9. Eguia, R. T. et al. A human coronavirus evolves antigenically to escape antibody immunity. PLoS pathogens 17, e1009453 (2021).

10. Chalkias, S. et al. A bivalent omicron-containing booster vaccine against covid-19. New Engl. J. Medicine 387, 1279–1291 (2022).

11. Tegally, H. et al. The evolving sars-cov-2 epidemic in africa: Insights from rapidly expanding genomic surveillance. Science 378, eabq5358 (2022).

12. Brito, A. F. et al. Global disparities in SARS-CoV-2 genomic surveillance. Nat. Commun. 13, 7003 (2022).

13. Khare, S. et al. Gisaid’s role in pandemic response. China CDC Wkly. 3, 1049 (2021).

14. Funk, T. et al. Characteristics of SARS-CoV-2 variants of concern b.1.1.7, b.1.351 or p.1: data from seven EU/EEA countries, weeks 38/2020 to 10/2021. Euro Surveill. 26 (2021).

15. Tegally, H. et al. Sixteen novel lineages of SARS-CoV-2 in south africa. Nat. Med. 27, 440–446 (2021).

16. Tegally, H. et al. Emergence of SARS-CoV-2 omicron lineages BA.4 and BA.5 in south africa. Nat. Med. 1–6 (2022).

17. Stockdale, J. E., Liu, P. & Colijn, C. The potential of genomics for infectious disease forecasting. Nat Microbiol 7, 1736–1743 (2022).

18. Huang, C. et al. Clinical features of patients infected with 2019 novel coronavirus in wuhan, china. Lancet 395, 497–506 (2020).

19. Hill, S., Perkins, M. & von Eije, K. Genomic sequencing of SARS-CoV-2. Tech. Rep., World Health Organization (2021).

20. Davies, N. G. et al. Estimated transmissibility and impact of SARS-CoV-2 lineage b.1.1.7 in england. Science 372 (2021).

21. Obermeyer, F. et al. Analysis of 6.4 million SARS-CoV-2 genomes identifies mutations associated with fitness. Science 376, 1327–1332 (2022).

22. Jankowiak, M., Obermeyer, F. H. & Lemieux, J. E. Inferring selection effects in sars-cov-2 with bayesian viral allele selection. PLoS genetics 18, e1010540 (2022).

23. Davies, N. G. et al. Estimated transmissibility and severity of novel SARS-CoV-2 variant of concern 202012/01 in england (2020).

24. Figgins, M. D. & Bedford, T. SARS-CoV-2 variant dynamics across US states show consistent differences in effective reproduction numbers (2021).

25. Vöhringer, H. S. et al. Genomic reconstruction of the sars-cov-2 epidemic across england from september 2020 to may 2021. medRxiv 2021–05 (2021).

26. CDC. Science brief: Emerging SARS-CoV-2 variants. https://www.cdc.gov/coronavirus/2019-ncov/science/science-briefs/scientific-brief-emerging-variants.html (2021). Accessed: 2021-4-16.

27. SARS-CoV-2 variants of concern and variants under investigation in england: Technical briefing 45. (2022).

28. SARS-CoV-2 variants of concern and variants under investigation in england: Technical briefing 43. (2022).

29. SARS-CoV-2 variants of concern and variants under investigation in england: Technical briefing 39. (2022).

30. Genomic surveillance of sars-cov-2 circulating in the united states. https://github.com/CDCgov/SARS-CoV-2_Genomic_Surveillance.

31. Borchering, R. K. et al. Modeling of future COVID-19 cases, hospitalizations, and deaths, by vaccination rates and nonpharmaceutical intervention scenarios - united states, April-September 2021. MMWR Morb. Mortal. Wkly. Rep. 70, 719–724 (2021).

32. Davies, N. G. et al. Increased hazard of death in community-tested cases of SARS-CoV-2 variant of concern 202012/01. medRxiv (2021).

33. Johnson, K. et al. Real-Time projections of SARS-CoV-2 b.1.1.7 variant in a university setting, texas, USA. Emerg. Infect. Dis. journal 27, 3188 (2021).

34. Cramer, E. Y. et al. The united states COVID-19 forecast hub dataset. Sci. Data 9, 1–15 (2022).

35. Kaiming Bi, Anass Bouchnita, Oluwaseun F. Egbelowo, Spencer Fox, Michael Lachmann, Lauren Ancel Meyers. Scenario projections for the spread of SARS-CoV-2 omicron BA.4 and BA.5 subvariants in the US and texas. (2022).

36. Reich, N. G. & Ray, E. L. Collaborative modeling key to improving outbreak response. Proc. Natl. Acad. Sci. U. S. A. 119, e2200703119 (2022).

37. SARS-CoV-2 genomics surveillance capacity map. https://www.finddx.org/covid-19/covid-19-genomic-surveillance/sars-cov-2-genomics-surveillance-capacity-map/ (2022). Accessed: 2022-12-20.

38. Tracking sars-cov-2 variants. https://www.who.int/activities/tracking-SARS-CoV-2-variants (2022). Accessed: 2023-3-10.

39. Fineberg, H. V. Pandemic preparedness and response–lessons from the H1N1 influenza of 2009. N. Engl. J. Med. 370, 1335–1342 (2014).

40. Lipsitch, M. & Santillana, M. Enhancing situational awareness to prevent infectious disease outbreaks from becoming catastrophic. In Inglesby, T. V. & Adalja, A. A. (eds.) Global Catastrophic Biological Risks, 59–74 (Springer International Publishing, Cham, 2019).

41. Kucharski, A. J., Hodcroft, E. B. & Kraemer, M. U. G. Sharing, synthesis and sustainability of data analysis for epidemic preparedness in europe. Lancet Reg Heal. Eur 9, 100215 (2021).

42. Hill, V. et al. Towards a global virus genomic surveillance network. Cell Host & Microbe (2023).

43. Campbell, F. et al. Increased transmissibility and global spread of sars-cov-2 variants of concern as at june 2021. Eurosurveillance 26, 2100509 (2021).

44. Du Plessis, L. et al. Establishment and lineage dynamics of the sars-cov-2 epidemic in the uk. Science 371, 708–712 (2021).

45. McCrone, J. T. et al. Context-specific emergence and growth of the SARS-CoV-2 delta variant. Nature 610, 154–160 (2022).

46. Johnson, K. E. et al. Real-time projections of sars-cov-2 b. 1.1. 7 variant in a university setting, texas, usa. Emerg. Infect. Dis. 27, 3188 (2021).

47. van Smeden, M. et al. No rationale for 1 variable per 10 events criterion for binary logistic regression analysis. BMC Med. Res. Methodol. 16, 163 (2016).

48. van Smeden, M. et al. Sample size for binary logistic prediction models: Beyond events per variable criteria. Stat. Methods Med. Res. 28, 2455–2474 (2019).

49. de Jong, V. M. T. et al. Sample size considerations and predictive performance of multinomial logistic prediction models. Stat. Med. 38, 1601–1619 (2019).

50. Rader, B. et al. Crowding and the shape of covid-19 epidemics. Nat. medicine 26, 1829–1834 (2020).

51. Taylor, B. P. & Hanage, W. P. A simple model of how heterogeneous disease transmission impacts the emergence of variants and their detection (2022).

52. Mlcochova, P. et al. Sars-cov-2 b. 1.617. 2 delta variant replication and immune evasion. Nature 599, 114–119 (2021).

53. Uriu, K. et al. Characterization of the immune resistance of severe acute respiratory syndrome coronavirus 2 mu variant and the robust immunity induced by mu infection. The J. Infect. Dis. 226, 1200–1203 (2022).

54. Kraemer, M. U. et al. Monitoring key epidemiological parameters of sars-cov-2 transmission. Nat. medicine 27, 1854–1855 (2021).

55. Tegally, H., Khan, K., Huber, C., De Oliveira, T. & Kraemer, M. U. Shifts in global mobility dictate the synchrony of sars-cov-2 epidemic waves. J. Travel. Medicine 29, taac134 (2022).

56. Balcan, D. et al. Multiscale mobility networks and the spatial spreading of infectious diseases. Proc. national academy sciences 106, 21484–21489 (2009).

57. Bajardi, P. et al. Human mobility networks, travel restrictions, and the global spread of 2009 h1n1 pandemic. PloS one 6, e16591 (2011).

58. pangoLEARN.

59. Hart, W. S. et al. Inference of the SARS-CoV-2 generation time using UK household data. Elife 11, e70767 (2022).

60. Hart, W. S. et al. Generation time of the alpha and delta SARS-CoV-2 variants: an epidemiological analysis. Lancet Infect. Dis. 22, 603–610 (2022).

61. Park, S. W. et al. The importance of the generation interval in investigating dynamics and control of new sars-cov-2 variants. J. The Royal Soc. Interface 19, 20220173 (2022).

62. Venables, W. N. & Ripley, B. D. Modern Applied Statistics with S (Springer, New York, 2002), fourth edn. ISBN 0-387-95457-0.

63. Elbe, S. & Buckland-Merrett, G. Data, disease and diplomacy: GISAID’s innovative contribution to global health. Glob Chall 1, 33–46 (2017).

64. Shu, Y. & McCauley, J. GISAID: Global initiative on sharing all influenza data - from vision to reality. Euro Surveill. 22 (2017).

65. Mathieu, E. et al. Coronavirus pandemic (COVID-19). Our World Data (2020).

