## Supplemental Information for "Leveraging global genomic sequencing data to estimate local variant dynamics"

January 2023

4 Data/code availability 35

5 EPI_SET identifier 35

### 1 Supplemental Methods

We describe a general method to estimate multi-strain dynamics and relative fitness advantages by partially pooling information across patches (geographic subunits, i.e. countries) and strains (Figure 2). This statistical approach leverages a hierarchical mixed-effects Bayesian framework. The model has two levels of hierarchy. In the first level, country-specific variant fitness advantages are structured such that the fitness advantage of a variant in one location informs the expected fitness advantages of variants in other locations to formalize the assumption that a variant's properties in one location are likely to be similar in another. In the second level, variants' mean fitness advantages, averaged over countries, consist of a shared (hierarchical) normal distribution. This approach shares information across variants to formalize the ecological assumption that most variants will be similarly fit to their recent ancestors and observing extreme deviations in fitness is uncommon. This assumption leads to shrinkage on extreme fitness advantage estimates for novel or otherwise infrequently observed variants for which we might otherwise overfit to noise in the data.

Our method addresses the twin issues of sparsity and bias by pooling the information from all sequencing done globally to build a picture of the relationships between variant dynamics and countries' individual characteristics (i.e., empirically observed correlations between variant fitness in specific countries, potentially driven by similarities in the immune landscape or contact patterns). This regularization approach is commonly used to stabilize estimates (e.g., with a ridge penalty or, equivalently, a standard normal prior) and, here, we extend this idea to partition sources of variance and apply multiple levels of regularization, verifying empirically that it reduces this transient bias (Figure 4). The resulting global landscape of variant fitness is then applied to individual countries, informing our understanding of variant dynamics in data-sparse regions, particularly in the early days after a variant's emergence (Fig. 3 and Fig. S3, S4, S5).

In the main text, we apply the model to SARS-CoV-2 sequences submitted to GISAID [5] up through July 1, 2022, focusing specifically on BA.4 and BA.5's global emergence.

#### 1.1 Data processing

Line list SARS-CoV-2 sequence metadata was accessed via the GISAID EpiCov database [5]. The findings of this study are based on metadata associated with 2,032,779 sequences available on GISAID up to July 1, 2022, via https://doi.org/10.55876/gis8.230118ka. Each row of the dataset contains information on the collection date, submission date, location of sample collection, and the assigned pango lineage [2] of the sequence submitted to the database. The country name in the location field is mapped to the corresponding three letter ISO country code. Sequence submissions missing complete date information (i.e. month and year instead of day month year) are excluded from the analysis and we assume this missingness is completely at random. Because pango lineage assignments are often delayed following initial submission, any sequences submitted on the reference date are excluded as they will still be labeled as "Unassigned" because they have not yet been assigned a lineage by GISAID's build of the Pangolin assignment tool [8]. Line list data was aggregated by country, collection date, and pango lineage to get daily counts of the number of sequences of each lineage observed in each country. In order to make the number of unique lineages tractable for model fitting and analysis, we manually set the lineages we are interested in tracking. During this time period, this corresponded to: BA.1, BA.2, BA.2.12.1, BA.4, and BA.5. All pango lineages except for those labeled as the variants we're manually tracking and WHO VOCs [14] were aggregated by the first number in their pango lineage assignment. For example, BA.5.1 would be assigned to the BA.5 lineage. We truncated the data to the past 90 days from the reference date for model fitting. Lineages with fewer than 50 observed sequences globally were aggregated into "other" along with the sequences labeled as "Unassigned". For visualization and model evaluation, we further collapse the pango lineage assignments into the "variants we're tracking", with all other pango lineages falling into the "other" category. Summary statistics (i.e. daily and weekly observed variant prevalence and standard errors) are calculated from this aggregation level, for comparison with model outputs.

#### 1.2 Hierarchical generalized linear modeling approach

We model the dynamics of competing variants of a directly transmitted infectious disease. This approach can be used generally for competing strains, but we use here the example of SARS-CoV-2 variants using sequence metadata from GISAID to produce estimates of relative variant growth rates and true proportion of total cases in a given country. Using a hierarchical generalized linear approach, the model shares information across countries and across variants to improve estimates of variant growth advantages and dynamics in settings with sparse sampling.

#### 1.3 Observation process

We consider the observed counts of lineages as drawn from a multinomial distribution. If we consider all sequences observed over the past 90 days (90 days ago $t = 0$; $t \in \mathbb{W}$ ), on day $t$ we observe $N_t$ sequences total ($N_t \in \mathbb{W}$), of which $n_{it}$ sequences are of variant $i$ ($n_{it} \in \mathbb{W}$; $N_t = \sum_i n_{it}$). Then the set of observed variants over the time period is $i_t \in \{i, ..., I\}$, where $I$ is the reference variant, which we set as the dominant lineage; the most frequently observed variant globally in the past 2 weeks. Although this quantity changes over time due to the emergence of mutations that warrant a new lineage designation, we consider it here to be both fixed and known (i.e. the number of unique Pango lineages known on day $t = 90$). The multinomial probability mass function (PMF) and its unknowns motivate the regression model formulation

$$Y_{ijt} \sim multinom(N_{jt}, p_{ijt}) \tag{1}$$

$$p_{ijt} = \frac{e^{\eta_{ijt}}}{\sum_{j=1}^{J} e^{\eta_{ijt}}} \tag{2}$$

$$\eta_{ijt} = \beta_{0ij} + \beta_{1ij} z_t \tag{3}$$

$$z_t = \frac{t - \mu_t}{\sigma_t} \tag{4}$$

Where $i$ indexes variants, $j$ indexes countries, $t$ indexes time, and $\mu^t$ and $\sigma^t$ describe the mean and standard deviation of the vector of timesteps where each timestep is a day (this transformation is discussed more in the Bayesian modeling approach subsection). The parameters $\beta_{0Ij} = \beta_{1Ij} = 0$ are fixed to ensure identifiability. The intercepts $\beta_{0ij}$ describe the initial variant prevalence on the scale of the linear predictor on day $t$. The slope coefficients ($\beta_{1ij}$) describe the difference in intrinsic growth rates for the variant in the numerator and the variant in the denominator (i.e. $\beta_{1ij} = r_{ij} - r_{Ij}$ where $r_I j$ is the intrinsic/Malthusian growth rate of the dominant variant in country $j$). However, this model does not produce an estimate of the actual intrinsic growth rates (i.e. $r_{ij}$ or $r_{Ij}$ ) because fitting $I$ cases would make the model overdetermined. This is why we refer to the $\beta_{1ij}$ terms as relative variant growth rates in the text. This formulation accounts for changes in sample size over time (i.e., changes in $N_t$) and, from these counts, estimates the expected proportion of the population made up of each variant. For interpretability, we present the global and country-specific estimates of the relative growth rates, $\beta_{1ij}$ and $\mu_{\beta_{1i}}$ as relative weekly fitness advantages using the relation:

$$f = e^{7\beta_{1ij}} - 1 \tag{5}$$

$$f = e^{7\mu_{\beta_{1i}}} - 1 \tag{6}$$

As described by Davies et al.[3]

#### 1.4 Hierarchical modeling structure

The hierarchical modeling approach pools information across variants and across countries, strengthening inference in settings with sparse information. This partial pooling addresses two forms of sparsity: variability in information available across variants because new variants have only been observed for a short period of time and systematic geographic variability in sequencing availability limiting the amount of information available in specific countries.

In the first layer of hierarchy, the model shares information across variants, making the assumption that most observed variants should be similar to each other. The model likelihood structures expected relative variant growth rates (i.e., $\mu_{\beta_{1i}}$ ) as drawn from a population distribution of growth rates:

$$\mu_{\beta_{1i}} \sim N(\mu_{\text{hierarchical}}, \sigma_{\text{hierarchical}}) \tag{7}$$

The parameter $\mu_{\text{hierarchical}}$ specifies the expected relative variant growth rate of any variant compared to the reference variant $I$, which we assign to be the dominant variant to increase interpretability and numerical stability. The parameter $\sigma_{\text{hierarchical}}$ specifies the expected amount of variability of mean relative variant growth rates around $\mu_{\text{hierarchical}}$. The parametric assumption of a normal distribution assumes that radically more or less fit variants than the bulk of the population is quite rare (i.e., the population is not leptokurtic). Additional details on model fitting are available in the Bayesian Model Approach subsection.

The model also shares information across countries, allowing variant dynamics in one country to inform estimates of variant dynamics in other countries. Individual countries have country-specific relative variant growth rates $\beta_{1ij}$, where $j$ indexes countries. The parameter $\mu_{\beta_{1i}}$ is a global estimate of relative variant fitness, pooled across countries. Systematic variation in growth rates across countries is learned empirically, expressed as a variance-covariance matrix $\Sigma$:

$$\beta_{0ij} \sim N(\mu_{\beta_0 i}, \sigma_{\beta_0 i}) \tag{8}$$

$$\begin{bmatrix} \beta_{11j} \\ \vdots \\ \beta_{1Ij} \end{bmatrix} \sim MVN(\begin{bmatrix} \mu_{\beta_{11}} \\ \vdots \\ \mu_{\beta_{1I}} \end{bmatrix}, \Sigma) \tag{9}$$

$$\Sigma = \begin{bmatrix} \sigma^2_{\beta_{11}} & & & & \\ \sigma_{\beta_{11}}\sigma_{\beta_{12}}\rho_{21} & \sigma^2_{\beta_{12}} & & & \\ \sigma_{\beta_{11}}\sigma_{\beta_{13}}\rho_{31} & \sigma_{\beta_{12}}\sigma_{\beta_{13}}\rho_{32} & \sigma^2_{\beta_{13}} & & \\ \vdots & & & \ddots & \\ \sigma_{\beta_{11}}\sigma_{\beta_{1I}}\rho_{I1} & \dots & & & \sigma^2_{\beta_{1I}} \end{bmatrix} = D(\sigma_{\beta_1})\Omega D(\sigma_{\beta_1}) \tag{10}$$

Country-specific vectors of relative variant growth rates are drawn from this multivariate normal distribution, with a shared vector of global means and empirically learned variability around this mean vector. This approach allows for stable estimates of variant dynamics in countries with limited sequencing capacity or when variants have only been observed in a limited subset of countries.

Systematic correlations between relative variant growth rates in countries (i.e.,$\beta_{1ij}$ ) is modeled by the variance-covariance matrix $\Sigma$ . This matrix can be decomposed into a correlation matrix $\Omega$ and a vector of independent lineage-specific variances $\sigma^2_{\beta_{1i}}$:

$$\Omega = \begin{bmatrix} 1 & & & & \\ \rho_{21} & 1 & & & \\ \rho_{31} & \rho_{32} & 1 & & \\ \vdots & & & \ddots & \\ \rho_{I1} & \dots & & & 1 \end{bmatrix}$$

$$D(\sigma^2_{\beta_1}) = \begin{bmatrix} \sigma^2_{\beta_{11}} & & & \\ & \sigma^2_{\beta_{12}} & & \\ & & \ddots & \\ & & & \sigma^2_{\beta_{1I}} \end{bmatrix}$$

This approach identifies systematic correlations between realizations of relative variant growth rates from this multivariate normal distribution. In other words, it identifies if realizations of variant $i$ in one country are systematically higher or lower when realizations of another variant are higher in that country. The variances of the $\beta_{1ij}$ realizations around the mean $\mu_{\beta_{1i}}$are independent, allowing differences in local dynamics to reflect changing variability across variants.

#### 1.5 Intercept structure

The model adds parametric structure to country-specific intercepts, enforcing the idea that variants are introduced to new countries at similar times. More formally, the variant-country intercept $\beta_{0ij}$ is drawn from a hierarchical student $t$ distribution:

$$\beta_{0ij} \sim t(\mu_{\beta_{0i}}, \sigma^2_{\beta_{0i}}, \nu = 2) \tag{11}$$

Unlike with the relative variant growth rates, there is no additional hierarchical structure on the means or variances of the intercepts $\beta_{0ij}$. The means $\mu_{\beta_{0i}}$ are left independent, to allow for independent and unstructured variant emergence. Likewise the $\sigma^2_{\beta_{0i}}$ parameters are left independent across variants. This allows for independent patterns of between-country variant invasion across variants and allows for different amounts of variability in prevalence on the initial day of the model. The parametric choice of a student $t$ distribution allows for substantial variability in timing between countries (i.e., kurtosis, heavy tails in the distribution).

#### 1.6 Model assumptions and limitations

All model estimates are based on SARS-CoV-2 sequences collected worldwide and shared by sequencing labs via the shared virus sequences from the GISAID

Initiative's EpiCov database. The modeling approach makes a number of simplifying assumptions to improve mathematical tractability. There are a number of limitations that may influence our estimates, both of variant fitness advantage globally and locally, as well as of variant prevalence dynamics.

Modeling assumptions include:

- The model assumes that sequences are randomly sampled within each geographic unit of infected individuals (e.g. country or state). In reality, different areas within countries often submit disproportionate numbers of sequences and these sequences may be submitted more quickly from one region than another. There is heterogeneity in sequencing coverage and speed even within a country or state, and thus, the observed variant proportions from sequenced samples may not be representative of the true variant proportions in the infected population.

- Sequencing labs do not prioritize particular samples for sequencing. If samples suspected to be from a particular variant are prioritized (e.g. if a variant is more severe, it may be more likely to be sequenced, or if a specimen shows a sign of a novel variant through a S-gene target failure, it may be selected for sequencing), the observed proportion of sequences from that variant will be artificially inflated due to the sampling process.

- All variants have the same generation interval. A pathogen's generation interval is defined as the time between infection and transmission. When the generation interval is shorter for one pathogen (or variant) than another, it can inflate the apparent growth rate [9].

- Relative transmission advantage of a variant within a country is constant over the 90 day period. Transmission advantage can change over time through a number of mechanisms, such as the buildup of immunity to a particular variant or the release of a new vaccine that induces a stronger immune response to a subset of the variants. The model does not account for any change, and instead assumes that the 90 day period is short enough that any change in advantage would be minor.

- Variant transmission is through a well-mixed population. Different parts of the population are more likely to come into contact with some parts than others (e.g., college students living in dorms likely see mostly other college students). If the population subsets that a variant is spreading within have higher contact rates or lower amounts of immunity than the general population, it can artificially inflate estimates of growth [13].

- Pango lineage assignment is perfect. The Pango Network defines new lineages, assigns a name, and builds and disseminates the PangoLEARN model to assign sequences to particular lineages [8]. Although lineage assignments are predicted from this model and can be incorrect, our modeling approach does not account for error in assignment and assumes all assignments are perfect. When lineage assignment is systematically incorrect, it can lead to biased estimates of fitness advantages.

- Collection dates and locations are correctly reported. Sequences shared by submitting labs via GISAID's EpiCoV database report the date and location of sample collection. The model used in this dashboard uses these dates and locations and assumes they are correctly reported.

- Relative fitness advantage is normally distributed across the population of variants (along with other parametric assumptions). This parametric assumption can shrink uncertain estimates toward the grand mean of variant fitness advantage. If the population is instead heavy-tailed or bimodal, this shrinkage could be inappropriate and lead to biased estimates.

- The effect of fitness advantage is linear on the scale of the linear predictor. The model assumes that the effect of fitness advantage on the growth rate is constant. However, this assumption could be incorrect due to changes in variant composition over time or immunity in the host population due to very quick growth. If incorrectly specified, model estimates would be biased.

- In addition, the model does not currently account for spatial structure in any form — neither in estimated variant prevalences nor in estimated intercepts. It does not take into account which countries are neighbors or in close spatial proximity (i.e., there is no spatial kernel). It also does not account for structure in variant invasion patterns due to, for example, airline mobility patterns. This approach likely loses some spatial information that could improve estimates, but also makes the model conceptually simpler and limits the amount of information needed to run it. The model structure also assumes that relative variant fitness advantage is constant over time and linear on the scale of the linear predictor. This assumption is plausible because of the short time frame over which the model is run — 90 days. Therefore, any systematic changes in the host population that would impact the fitness of particular variants is unlikely to be large enough to lead to large changes in fitness advantage.

These simplifying assumptions allow for this modeling approach to be feasible on the finite amount of available information about variant dynamics. Although the model is constrained by these assumptions, it is able to flexibly fit the available data, even for countries with limited sequencing capacity. Most of these biases affect the estimates for recent time windows with only partially complete sequencing, and as more sequences get submitted corresponding to a particular time window, these potential biases tend to be less apparent. What this means is that recent estimates of variant prevalence can be less reliable due to the incomplete nature of the data and that estimates of novel emerging variants may be biased and are subject to change as the data becomes more complete and the variant is detected in different populations with different immune landscapes.

#### 1.7 Bayesian modeling approach

We fit this model with a fully Bayesian approach to allow for flexible numerical sampling and directly interpretable parameters. We place informative priors on model hyperparameters:

$$\begin{aligned}
\mu_{\beta_{0i}} &\sim t(-5, 5, 3) \\
\sigma_{\beta_{0i}} &\sim N^{+}(2, 1) \\
\Omega &\sim LKJ(2) \\
\sigma_{\beta_{1i}} &\sim N^{+}(0.5, 2) \\
\mu_{\text{hierarchical}} &\sim N(-1, 0.5) \\
\sigma_{\text{hierarchical}} &\sim N(1, 0.1)
\end{aligned}$$

These informative prior distributions are derived from the posterior of a previous, independent model fit. This model was fit to a subset of European countries for a period corresponding to November and December of 2021 with non-informative priors. From that model fit, we use the first and second moments from these marginal posterior distributions for hyperparameters as weakly informative prior distributions except for the prior on $\Omega$ and the degrees of freedom on $\mu_{\beta_{0i}}$. These priors are set to general weakly informative priors as recommended by the Stan software creators, both for numerical simplicity and to allow for different correlations because of the different subsets of variants observed [12]. The time period for the original model fit used to develop the prior distributions does not overlap with any of the observed time points in any of the model fits interpreted here and the set of observed variants and the dominant variant are both different, suggesting that the priors provide reasonable scaling for the numerical sampler without being overfit to the specific time period or variants.

For computational and modeling tractability, the time covariate is centered and scaled:

$$z_t = \frac{t - \mu_t}{\sigma_t} \tag{12}$$

Where $\mu_t$ and $\sigma_t$ are the mean and standard deviation of the vector of timesteps, with one day as one timestep. As a result, the priors on $\mu_{\text{hierarchical}}$, $\sigma_{\text{hierarchical}}$, and $\sigma_{\beta_{1i}}$ are on the scale of one standard deviation in scaled time (i.e. going from 0 to 1 on this scale is about 26 days). All hierarchical distributions are in a non-centered parameterization except for the distribution of $\mu_{\beta_1 i}$ which is in the centered parameterization.

The model is fit using Hamiltonian Monte Carlo (HMC) with the No-U-Turn sampler in CmdStan v2.29.2 [1]. Using HMC instead of a more approximate variational approach allows for full characterization of posterior uncertainty. We run the model with 4 parallel chains with 2500 warmup iterations and 500 sampling iterations per chain for a total of 2000 sampling iterations. For all fits, the Gelman-rubin split $\hat{r}$ statistic was less than 1.01, no samples hit the maximum treedepth of 10, there were no divergences, and E-BFMI is above 0.3, suggesting that the numerical sampler converged and was able to perform unbiased sampling.

### 2 Retrospective validation of country-specific variant prevalence projections

#### 2.1 Processing and fitting of historical datasets

Line-list sequence data from GISAID was accessed on the following dates, which we refer to as "reference" dates: April 30th, 2022, May 16th, 2022, May 27th, 2022, June 4th, 2022, June 27th, 2022 and July 1st, 2022. A consensus set of lineages was found by identifying all lineages that exceed the global threshold of 50 or more observed sequences in the past 90 days across any of the 6 reference datasets. These datasets were each processed as described in Section 1.1. We fit the multicountry model to each reference dataset independently using all sequences collected within the 90 days preceding the reference date and the same consensus set of lineages to be estimated. We define the calibration period as the period over which any sequences were collected and had been submitted by the reference date globally in GISAID, with the forecast period defined by any days without any observed sequences globally. To test the model's predictive power, we forecast 21 days out and compared this to any data observed by the last reference dataset from July 1st, 2022. The model returns the mean variant prevalence estimate for both the 90 day time period of calibration and the 21 day forecast.

#### 2.2 Estimation model comparison: Multicountry vs. Single country stability over time

For each reference dataset, we compare variant prevalence estimations from the multicountry model with estimations from single country multinomial models using the nnet package version 7.3.17 in R [11]. The nnet package returns the maximum likelihood estimation (MLE) of variant fitness advantages and mean variant prevalences. In order to get an estimate of the uncertainty of these outputs, non-parametric bootstrapping with replacement was performed, generating 100 boot-strapped datasets with time points randomly sampled with replacement from the true data. Each boot-strapped dataset was fit using the nnet multinom function.

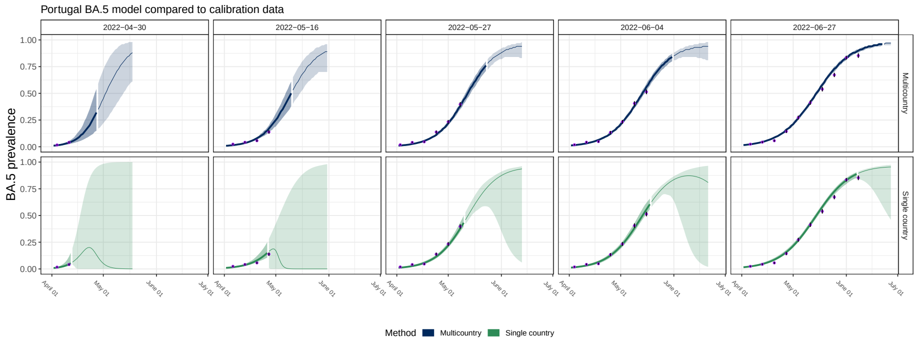

Figure S1: Multicountry (top) and single country (bottom) model estimated variant prevalence compared to the data observed at that time in Portugal for BA.5. Each column represents the date that the models were run and estimates were made (reference date). Shading indicates the calibration period (darker) and the forecast period (lighter). Line indicates the median projection. Purple dots indicate the weekly average observed prevalence of BA.5 as of that reference date, with bands indicating the standard error

Figure S1 shows the model estimated BA.5 proportions in Portugal compared to, in this instance, the observed variant proportions as of that reference date (purple points), for the multicountry model and the single country model. In the main text in Figure 4, we compare the model estimates to the observed variant proportions as of July 1st, 2022. These differ because of the delay from specimen collection to sequence submission, resulting in backfilling of prior proportion estimates as new sequences get added.

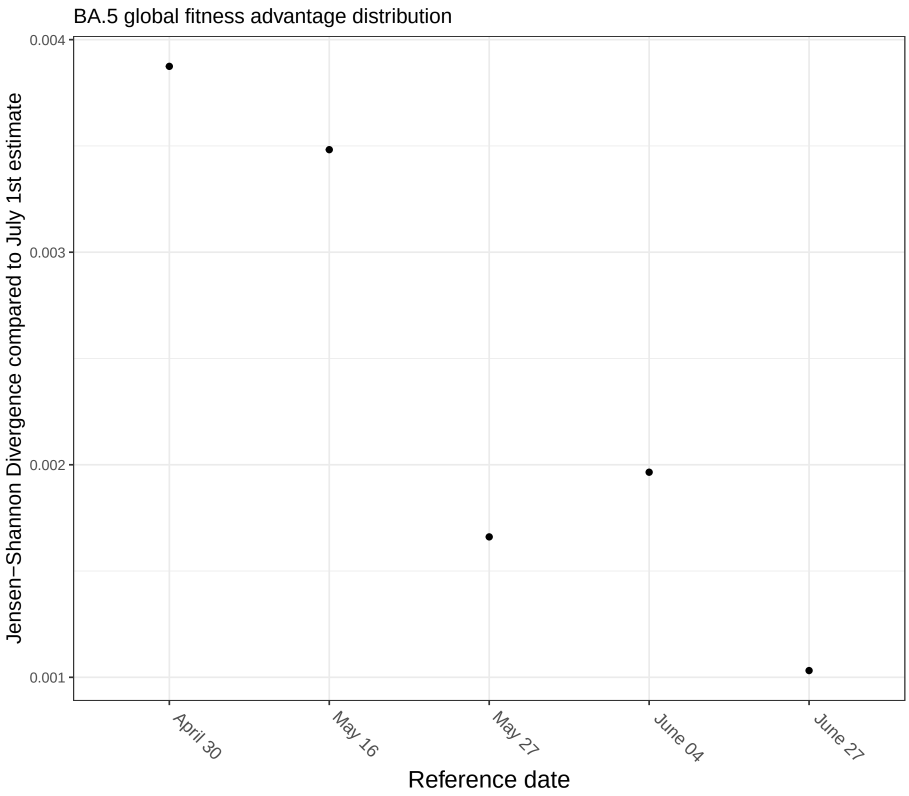

Figure S2: Evaluating the change in the BA.5 global fitness advantage posterior distribution using the Jensen-Shannon Divergence. Each point represents the Jensen-Shannon divergence between BA.5's global fitness advantage distribution estimate at the reference date of interest compared to that estimated at the last time point, July 1st 2022.

In Figure S2, we evaluate the stability of BA.5's global fitness advantage distribution over time (Figure 4B) using the Jensen-Shannon (JS) divergence as a measure for the difference between distributions, with the distribution from each reference date being compared to the estimated distribution from the data on July 1st, 2022. While the JS divergence does decrease over time as we would expect, the JS divergence is relatively low throughout the whole time period.

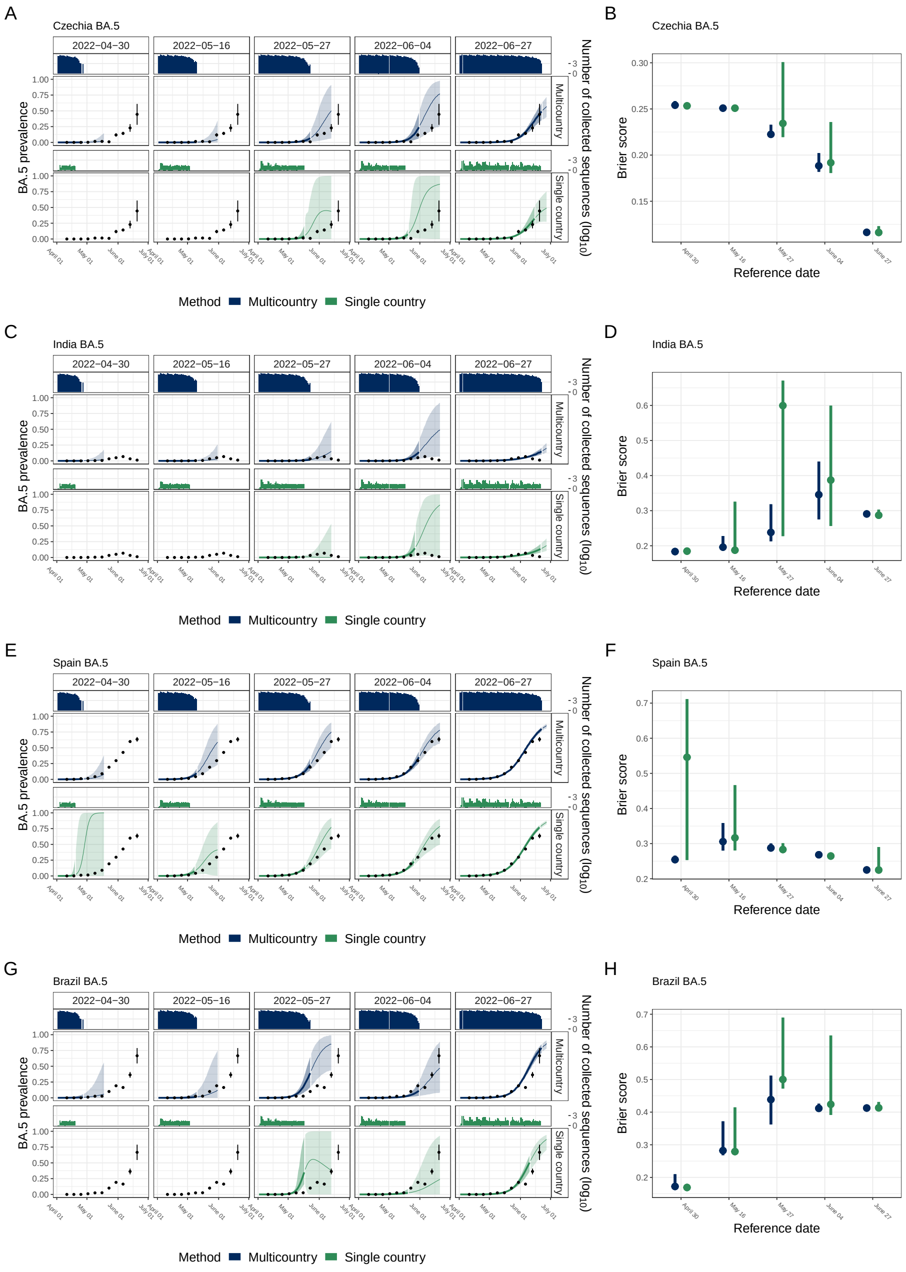

Figure S3: Retrospective analysis of estimated BA.5 variant prevalence dynamics. Model estimated BA.5 prevalence in Czechia, India, Spain, and Brazil from 5 reference dates (columns) and two models (rows) (A, C, E, &G). Color indicates the model and bar plots indicate the number of BA.5 sequences available to the model. Uncertainty bands represent the 95% credible interval of the multicountry model (blue) and the 95% confidence interval from the non-parametric bootstrap (green), with shading indicating the calibration period (darker) and the forecasting period (lighter). Black points indicate the observed variant prevalence as of July 1st, 2022, with error bars indicating the standard deviation. (B, D, F, &H) Brier score evaluating predictive accuracy of estimated BA.5 prevalence compared to the observed prevalence on July 1st, 2022.

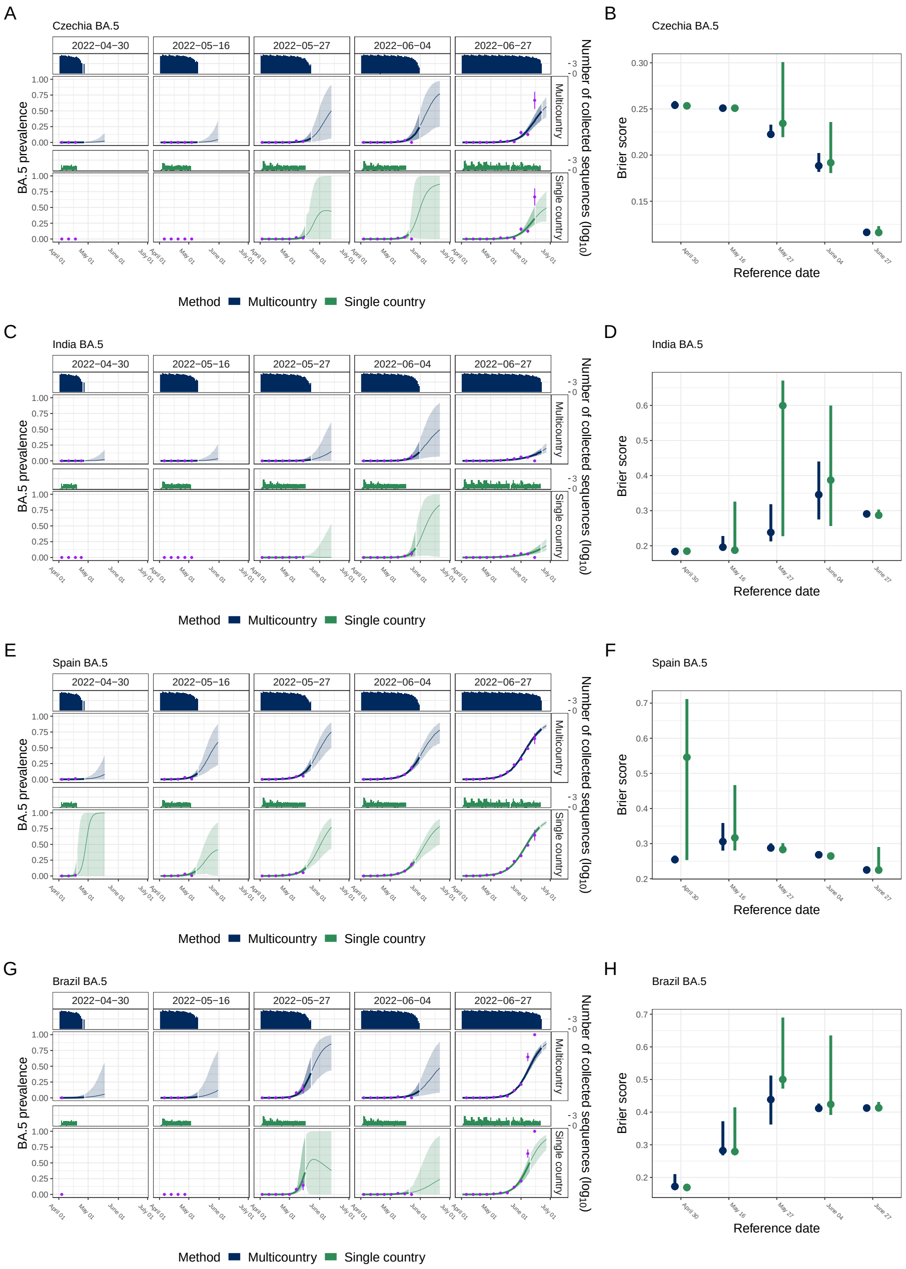

Figure S4: Examples of retrospective analysis of BA.5 prevalence dynamics, compared to the observed data as of that reference date. (A, C, E, &G) Each column represents the date that the models were run and estimates were made (reference date). Shading indicates the calibration period (darker) and the forecast period (lighter). Line indicates the median projection. Purple dots indicate the weekly average observed prevalence of BA.5 as of that reference date, with bands indicating the standard error.(B, D, F, &H) Brier score evaluating predictive accuracy of estimated BA.5 prevalence in Czechia, India, Spain, and Brazil compared to the observed prevalence on July 1st, 2022 as described in Section 2.3. Points indicate the mean Brier score, bars indicate the 95% credible interval on the Brier score from the multicountry model, and the 95th percentiles of the non-parametric bootstrapped estimates for the single country model.

We performed this retrospective validation analysis across all countries that had sequences collected within 90 days of the reference date and shared with GISAID. In Figure 4 A & C in the main text, we use Portugal's BA.5 emergence as a country-specific example of where the multicountry model can improve model estimates of variant dynamics. To demonstrate the generalizability across countries of this phenomenon, in Figure 4 D & E in the main text, we show the fitness advantage estimates from a subset of 9 countries, for the 5 reference dates and 4 key variants that were emerging at the time. Here, in Figure S3, we show results analogous to what is shown in Figure 4 A and C in the main text, but for 4 other countries with different BA.5 emergence timelines and sequencing capacity. In Figure S4, we compare the model estimated variant proportions at each reference date with the data they were fit to at that time. This reveals what we know to be true and difficult about real-time variant dynamics estimates: (1) the observed data do shift significantly over time as sequences from recent collection dates get backfilled and (2) the multicountry method is able to pull information from other country predictions while the single country model is forced to only rely on that minimal early data, and thus is overfitting to the observations.

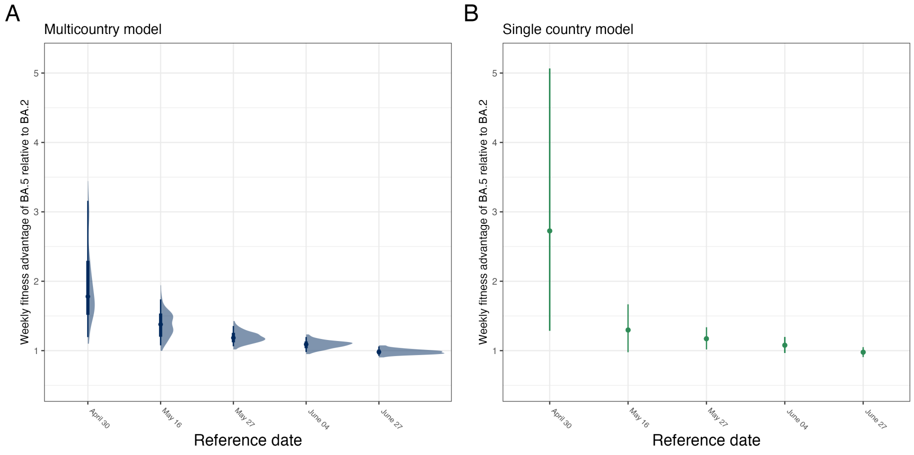

Figure S5: Multicountry (A) and single country (B) estimates of the weekly fitness advantage of BA.5 at each reference date. A. For the multicountry model, the posterior distribution of the estimated weekly fitness advantage is shown as of each reference date. Points indicate the median, bands indicate the 95th and 75th percentiles of the posterior distribution. B. Single country model estimates of the estimated weekly fitness advantage is shown as of each reference date. Points indicate the estimated fitness advantage applied to the observed data, bands indicate the 95% confidence intervals on the single country model using the normal approximation.

In Figure S5, we compare the estimates of the Portugal-specific weekly fitness advantage from the multicountry model and the single country model in order to compare the stability of the estimated variant fitness advantage in a particular region. For the multicountry model, we see that the early estimates do shift over time, however the median estimate is much closer to the later estimate than is the case for the single country model. We hypothesize that the reason for this early overestimate in the single country model could be due to factors such as demographic stochasticity, sampling bias, and overfitting to noisy data when data is sparse. In the main text, we show that the global estimated variant fitness advantage of BA.5, while initially more uncertain, remains relatively stable over the 5 reference dates (Main text, Fig 4B).

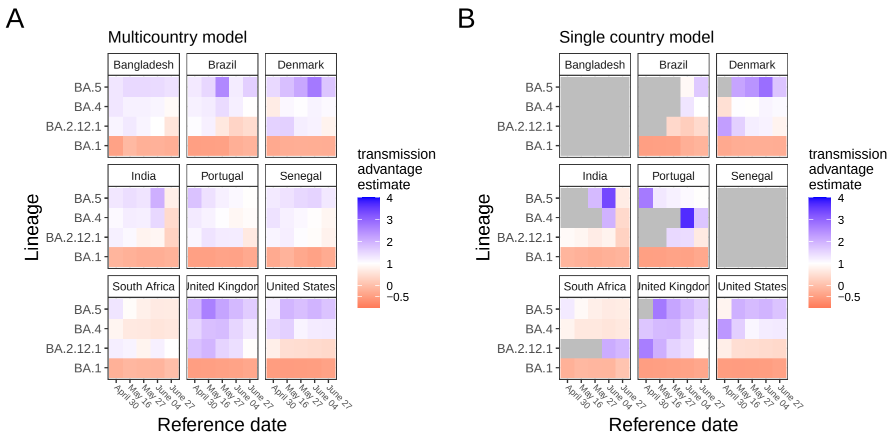

Figure S6: Comparison of the country-specific fitness advantage estimates across reference date and countries. A. Multicountry model estimates across reference dates show relative stability, even at early time points for BA.4 and BA.5. B. Single country model estimates across reference dates show the greater decline in estimated fitness advantage of emerging variants such as BA.5 over time (i.e. in the UK and Portugal), and overall less stability than in the multicountry model. Gray indicates time periods where there was insufficient data for model convergence within the country for that variant (a problem we don't see in the multicountry model because it leverages data for that variant from other countries).

In Figure S6, we show across a range of countries the estimated country-specific transmission advantage for the multicountry model (Figure S6A) and the single country model (Figure S6B). What we see is that there is greater fluctuation over time (left to right in each heatmap) in the estimates in the single country model compared to the relative stability of the estimates from the multicountry model. Likewise, we are able to make estimates of country-specific fitness advantages for all country-variant combinations in the multicountry model, whereas in the single country model, if that variant has not been observed in that country or there is not sufficient data, the model is unable to converge (gray gaps).

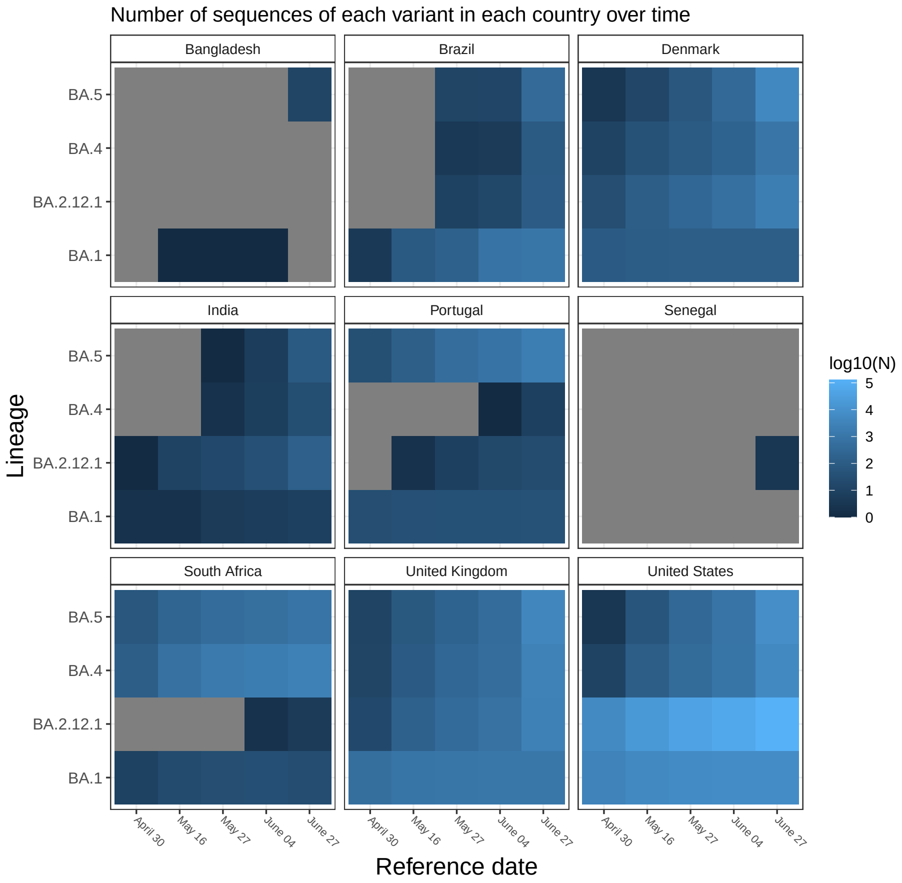

Figure S7: Number of sequences of each variant observed in each country by each reference date. Gray boxes indicate that that particular variant had not been observed in that country by that reference date. In these instances, single country estimation methods are unable to estimate that variant's fitness advantage within the country.

In Figure S7 we show the number of sequences of a particular variant in each country for the 5 reference dates. We see that for newly emerging variants such as BA.4, BA.5, and BA.2.12.1, a number of countries had yet to observe any sequences of that variant at the earlier reference dates, rendering it impossible to estimate using a single country method, the variants fitness advantage in that country (resulting in corresponding gray boxes in Figure S6B).

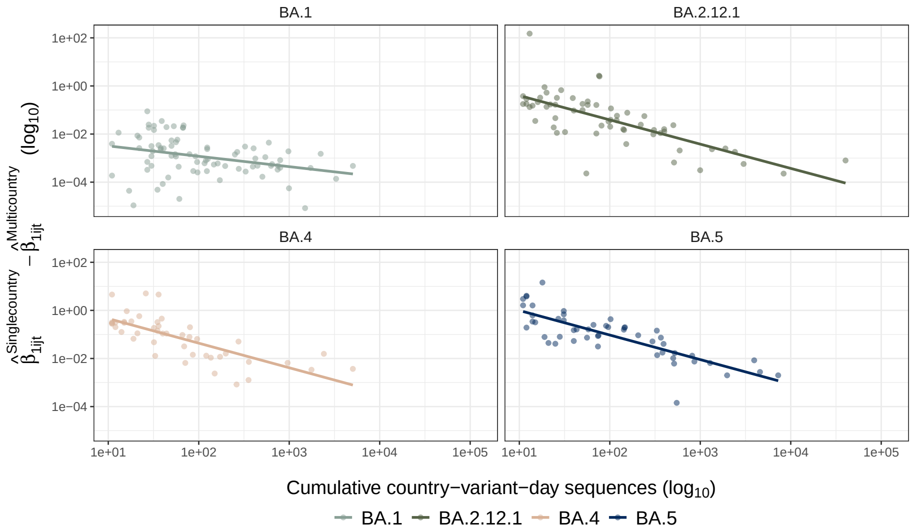

Figure S8: Difference between country-specific fitness advantage from the multicountry model ($\hat{\beta}_{ijt}^{\text{multicountry}}$) and single-country models ($\hat{\beta}_{ijt}^{\text{multicountry}}$). The log differences in the two estimates (y-axis) are plotted against log cumulative sequences of a variant observed in a country on a day (x axis).

In Figure S8, we compare the difference between the multicountry fitness advantage estimate and the single country fitness advantage estimate for four key variants as a function of the number of observed sequences of that variant in that country up until the time the estimation was made. The estimated coefficients from the multicountry model are consistently lower at low sequence counts than those estimated by single country maximum likelihood multinomial regressions. The maximum likelihood estimate coefficients are both higher initially and decline more slowly with the increase in sample size than those from the multicountry model. We note, however, that the comparison in terms of sample size is not one-to-one because the multicountry model uses all global sequences, while the single country is constrained to total within-country sequences (x axis values). Estimates from models fit on country-variant-days with fewer than 10 cumulative sequences are discarded to remove extreme outliers.

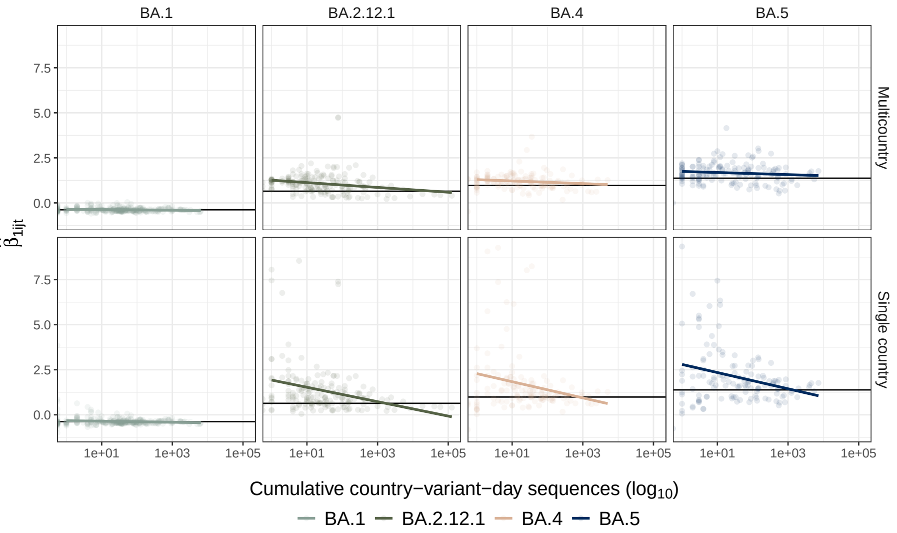

Figure S9: Estimated country-specific fitness advantage ($\hat{\beta}_{ijt}^{\text{multicountry}}$). Horizontal lines are the average of the estimates from the top 10% of cumulative sequences for the variant-lineage bin to visually approximate the asymptote. Estimated fitness advantages greater than 10 are removed for visual clarity, but all such estimates are from the MLE model and would further exaggerate the contrast (n = 44).

In Figure S9, we compare the estimated weekly variant fitness advantage for 4 key variants for the multicountry model (top) and the single country model (bottom) versus the number of observed sequences of that variant in that country up until the time the estimation was made. Maximum likelihood estimates are higher initially and converge more slowly in the single country model than in the multicountry model estimates, highlighting the improved stability of the multicountry model estimates. This is particularly evident in the case of BA.5 (right column) where multicountry estimates are relatively stable while the single country estimates decline significantly as the number of sequences increases.

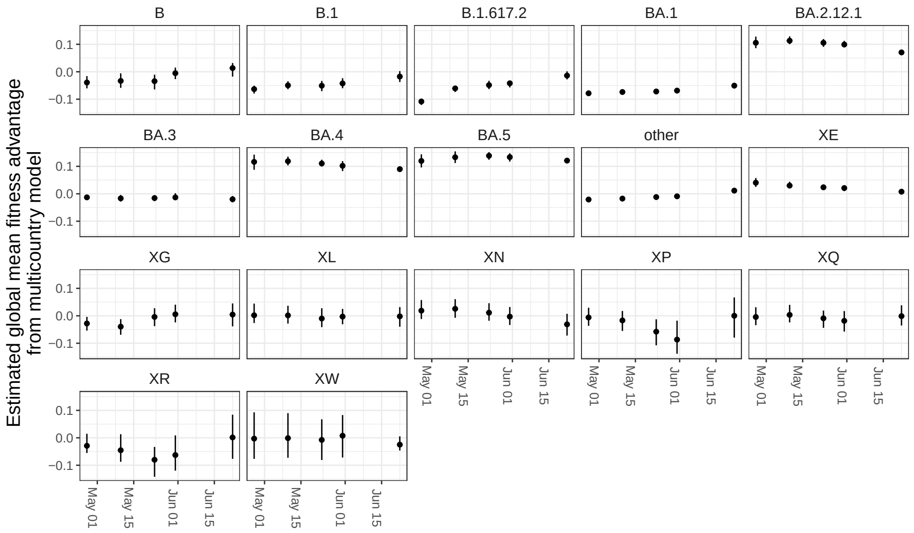

Figure S10: Estimated global fitness advantages from multicountry model over time. Estimated global means of variant fitness advantages estimated across the 5 reference dates.

In Figure S10, when we look at the estimates of global variant fitness advantage over time we see that estimates are stable with no systematic trend across variants. The lack of trend systematic trend across variants suggests that this estimator is appropriate to evaluate the risk posed by an emerging variant.

#### 2.3 Evaluation of model estimates using the Brier score

For both the multicountry and the MLE model, we used 100 draws from the distributions of lineage prevalence estimates to evaluate the accuracy of the model predictions compared to the observed daily lineage prevalence from the July 1st, 2022 reference dataset. For countries that observed no sequences of a particular lineage in the consensus dataset during the 90 day time window, the MLE estimation model does not estimate a prevalence for that lineage. To enable a fair comparison of the two model outputs at the country-level, we collapse all prevalence estimations from the multicountry model for these unobserved lineages into "other" for that country. We use the Brier score to evaluate the accuracy of each draw of the model output. The Brier score is calculated at the country-level for both the calibration period and the forecast period, and the two combined. Because we have a distribution of probabilistic predictions from our model output (variant proportions over time), we get a distribution of the brier score at each reference time point as a result of the evaluation process.

To estimate the Brier score for each country and reference dataset from the multinomial model output, we compare the mean estimated probability that a sequence is the $i$th lineage at the $t$th time, ($\hat{p}_{t,i}$), to the observed binary outcome $\Psi_{i,t,k}$ of whether the $k$th sequence on the $t$th day is the $i$th lineage.

$$BS = \frac{1}{\sum_{t=1}^{\tau} N_t} \sum_{t=1}^{\tau} \sum_{i=1}^{l} \sum_{k=1}^{N} (\hat{p}_{t,i} - \Psi_{i,t,k})^2$$

Where $N_t$ is the total number of sequences collected on the $t$th day, $l$ is the number of possible outcomes, in this case, the number of unique lineages estimated, and $\tau$ is the number of time points in the time window of interest. $\tau$ will depend on whether we are computing the Brier score for the calibration period, the forecast period, or the combination of the two.

For computational efficiency, instead of lining up the set of 1s and 0s for each lineage for each sequence collected each day, we can use the fact that and $\sum \Psi$ and $\sum \Psi^2$ are sufficient statistics for $\sum(p - \Psi)^2$ to evaluate the Brier score for each lineage at each time point. If we expand the above equation:

$$BS = \frac{1}{\sum_{t=1}^{\tau} N_t} \sum_{t=1}^{\tau} \sum_{i=1}^{l} \sum_{k=1}^{N} (\hat{p}_{t,i}^2 - 2\hat{p}_{t,i}\Psi_{i,t,k} + \Psi_{i,t,k}^2)$$

Which is equivalent to:

$$BS = \frac{1}{\sum_{t=1}^{\tau} N_t} \sum_{t=1}^{\tau} \sum_{i=1}^{l} (N\hat{p}_{t,i}^2 - 2\hat{p}_{t,i} \sum_{k=1}^{N} \Psi_{i,t,k} + \sum_{k=1}^{N} \Psi_{i,t,k}^2)$$

Replacing the sum of the binary (1,0) $\Psi_{i,t,k}$ with the number of sequences of each lineage, $n_{i,t}$ , as such, $\sum \Psi_{i,t,k}^2 = n_{i,t}$ we can write the Brier score in terms of the number of observed sequences of the $i$th lineage ($n_{i,t}$) and the total number of sequences, $N_t$ collected each day.

$$BS = \frac{1}{\sum_{t=1}^{\tau} N_t} \sum_{t=1}^{\tau} \sum_{i=1}^{l} (N_t\hat{p}_{t,i}^2 - 2\hat{p}_{t,i}n_{i,t} + n_{i,t})$$

The Brier score is calculated for each draw from the multinomial output for each country and each time period evaluated.

#### 2.4 Comparison to spline-based approach

In the main text, we compare the stability and efficiency of the growth rate estimates and fitted variant proportions from the hierarchical multicountry multinomial modeling approach developed here ("multicountry model") to that of separate multinomial models fit to the data from single countries ("single country model"). We chose to compare against this simple benchmark because it is common and because more sophisticated derivatives of this approach are used to inform situational awareness for public health (e.g., the US CDC's nowcast) [10].

As a simple baseline, it highlights two of the benefits of the additional structure from a hierarchical modeling approach: increased stability of estimates of novel variants and improved model convergence and stability for estimates in locations with sparse sequencing.

However, multinomial regression over a longer time period with data from one or several regions has also been used to estimate variant dynamics (see e.g., [3, 7, 4]). In this approach, relative variant fitness is modeled as a smooth function of time, rather than a linear relationship. Relative variant fitness can change if, for example, immunity to one variant built up in a population sufficiently to shift the advantage towards other variants with less prior immunity. By allowing for relative variant fitness to change as a function of time this mechanistic process can be reflected in the model. This modeling approach can be implemented using a spline of time [17, 3, 15] and can be a particularly effective tool when describing a past emergence of a new dominant variant and the resulting global changes in variant dynamics over a longer period of time. However, spline models (particularly unpenalized spline models) can perform poorly when data are limited or when extrapolating beyond the edges of the available data [17]. In contrast, we make the explicit assumption in the method presented in this manuscript that within a relatively short time window (e.g. 90 days) relative variant fitness advantage estimates are time-invariant. This choice allows us to perform the historical validation of our estimates, by assuming the estimates ($\beta_{ij}$ and $\mu_{\beta_i}$) should be constant over this short time period.

We compare our proposed approach to a spline-modeling approach from [16]:

$$y_{ijt} \sim multinom(N_{jt}, p_{ijt})$$

$$p_{ijt} = \frac{e^{\eta_{ijt}}}{\sum_i e^{\eta_{ijt}}}$$

$$\eta_{ijt} = A_{ij}\theta + f(i,t) + f(i,t,j \in \text{region})$$

$$f(\dot{}) = \sum_{m=1}^{2} b_m w_m$$

$$i \in \{\text{variants}\}, j \in \{\text{countries}\}, t \in \{\text{time}\}$$

Where $A_{ij}$ is a matrix with the intercept and indicator variables pertaining to a particular country-variant combination, $\theta$ is the associated vector of coefficients, $f(i,t)$ is a smooth natural spline with two knots describing the smooth trend in relative fitness of a particular variant over time, and $f(i,t,j \in \text{region})$ is a smooth natural spline with two knots describing the smooth trend in deviation of relative fitness of a particular variant in a particular region over time from the global smooth trend. Note, that this formulation does not have a fixed effect for region, only ones for country (the coefficients in the vector $\theta$). Because this model has multiple covariates with respect to time, one needs to take the marginal effect to get a one-number summary of the change with respect to time. In particular, one can estimate the marginal effect of time for fixed value of time, $t = t^*$, averaged over countries $E[\frac{\partial \eta_{ijk}}{\partial t} | t = t^*, i = i^*]$, and one can use the contrasts (i.e., the differences) of these marginal effects to estimate the relative growth rates of two variants: $\delta(i = i^*, i^+, t = t^*) = E[\frac{\partial \eta_{ij}}{\partial t} | t = t^*, i = i^*] - E[\frac{\partial \eta_{ij}}{\partial t} | t = t^*, i = i^+]$ for some arbitrary time, $t^*$, and some arbitrary variants $i$ and $i^+$ where $i^+ \neq i^*$. We will assume $i^+$ is the dominant variant and leave it out of subseqeunt notation for clarity. In the multicountry model, one can also calculate the marginal effect with respect to time: $E[\frac{\partial \eta_{ijt}}{\partial t} | i = i^*] = E[\beta_{1ij} | i = i^*] = \mu_{\beta_{1i}}$ – in other words, the marginal effect is the quantity we evaluate in the main text.

The parameter $\delta(i, t)$ is a time-dependent growth rate – it refers to the difference between the expectations of the realized, instantaneous growth rates of two variants on the scale of the linear predictor. In contrast, the parameter $\mu_{\beta_{1i}}$ estimated from the model proposed in the main text is the expectation of the differences between the time-invariant growth rates for a particular variant and the dominant variant $d$ in a particular country: $\mu_{\beta_{1i}} = E[\beta_{1ij} | I = i] = E[r_{ij} - r_{dj} | I = i]$.

While both $\delta(i, t)$ from this model and $\mu_{\beta_{1i}}$ from the hierarchical multinomial model we present in the main text are measures of the relative growth of a variant, they have a number of important differences and are not directly comparable. The parameter $\delta(i, t)$ measures the difference between the expected values of the instantaneous growth rates of two variants, averaged over countries and regions. The parameter $\mu_{\beta_{1i}}$ measures the expected difference between two time-invariant (i.e., malthusian) growth rates of two variants, averaged over countries. Therefore, these parameters measure different statistical quantities and are not directly comparable. These differences are practical: the difference of expectations is not equal to the expectation of the difference of random variables, with only the latter taking into account the joint distribution of the variables. Notably, the expectation of the difference in growth rates is a measure of central tendency describing whether a variant is taking over within countries, while the difference in expectations is not guaranteed to be a description of any particular population (e.g., Simpson's paradox could lead to the illusion of dominance). However, these differences are also conceptual: the difference in time-invariant growth rates refers to the difference between idealized malthusian growth rates – not instantaneous growth rates. Furthermore, these estimands are also not comparable because the covariates of a multinomial regression model are noncollapsible and so the average of coefficients corresponding to the subgroups (i.e., for time in this spline model) does not equal the single coefficient corresponding to time in the hierarchical model. For this reason, these one-number summaries of variant growth rates from these models cannot be directly compared.

Rather than directly compare coefficients from the fits from the spline-based multinomial model to the hierarchical model that we develop here, we evaluate the performance of the spline-based model by comparing the raw predictive accuracy each model. We also evaluate the stability of the estimated marginal relative growth rates (i.e., $\delta(i, t)$) across reference dates of the spline modeling approach, similar to how we evaluate the multicountry model estimates over time in the main text. We do not directly compare stability or the parameter magnitudes for the reasons listed above. We find that estimated relative fitness for the emerging BA.5 variant varies substantially over the reference dates (Section 2.4). For the comparison between the BA.5 and BA.2 variants, the standard errors for the contrast substantially underpredict the variation in estimates over the reference dates. Despite this instability, we find the two models have very similar predictive performance, with the spline-based fits generating similar brier scores to the hierarchical model (Figure S12). However, we note that the spline-based model can display substantial predictive inaccuracy in low sample-size settings (e.g., Morocco) (Figure S12). We also present fitted and predicted probabilities for the BA.1, BA.2, BA.4, BA.5, BA.2.12.1, and all other variants (Figures S13 to S18), but we are unable to estimate standard errors for the predictions because the normal approximation to the standard error would be inappropriate and the bootstrap standard errors would be computationally infeasible (one fit takes $\approx 6$ hours on these datasets).

Based on these results, we find that the spline-based model performs quite well fitting to the observed data and at nowcasting. In settings with lower computational cost (i.e., monthly or even weekly), we would advocate for its use with bootstrap standard errors provided the sample size is reasonable in all spatial units. However, we would caution against direct interpretation of the growth rates with respect to time for the reasons discussed above.

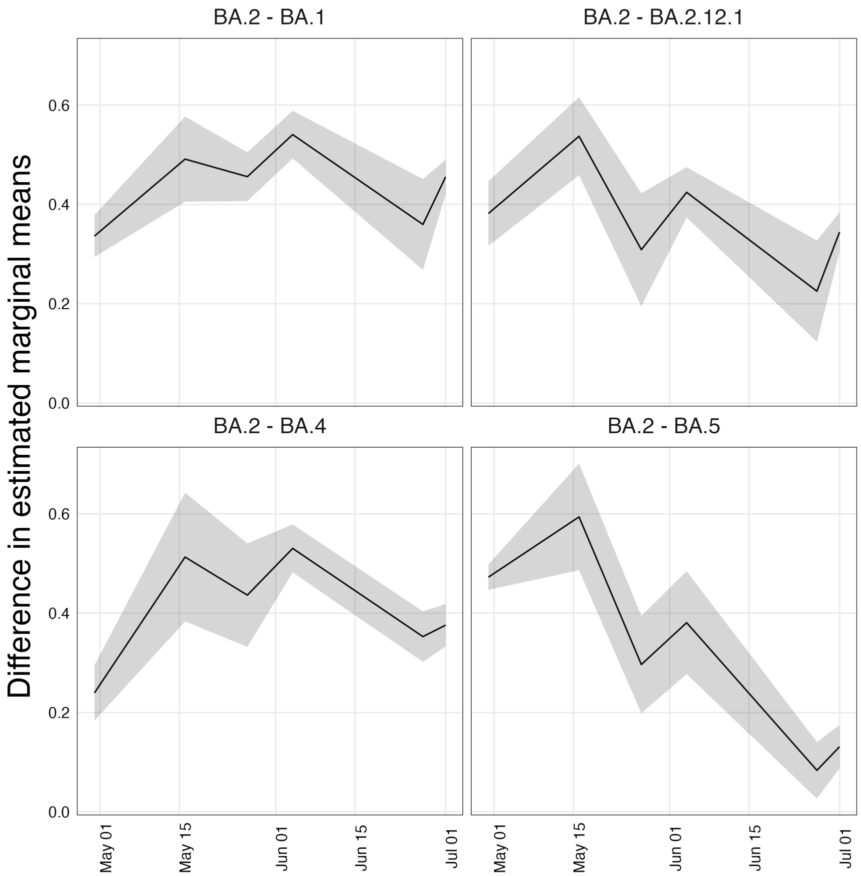

Figure S11: The contrasts between the expected marginal means for variants for interest and the dominant BA.2 variant across reference dates. The spline modeling approach was fit to each reference date dataset, with datasets for each reference date prepared as described in Section 2. At each timepoint the contrasts between the expected marginal means was computed using ‘emmeans‘ on the scale of the linear predictor averaging over countries and regions using a reference grid specifying the BA.1, BA.2, BA.2.12.1, BA.5, and BA.5 lineages and a timepoint 20 days past the last day in the reference dataset [6].

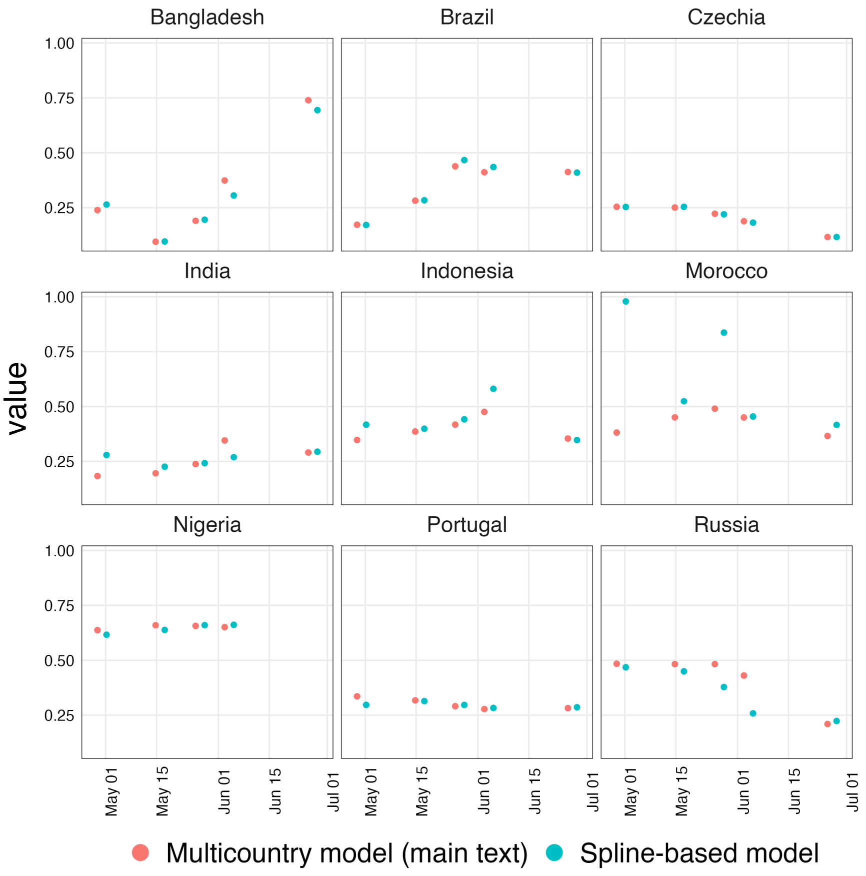

Figure S12: Comparison of the brier score from the spline model to that from the multicountry model. Both modeling approaches were fit to each of the reference dates, with datasets for each reference date prepared as described in Section 2. For each modeling approach, the brier score was computed for each model fit to a reference date using the dataset from the final reference date (2022-07-01) for the 90 period that the model was fit to and the 20 day forecast. All lineages that were not BA.1, BA.2, BA.2.12.1, BA.5, and BA.5 were aggregated to "other" before computing the brier score.

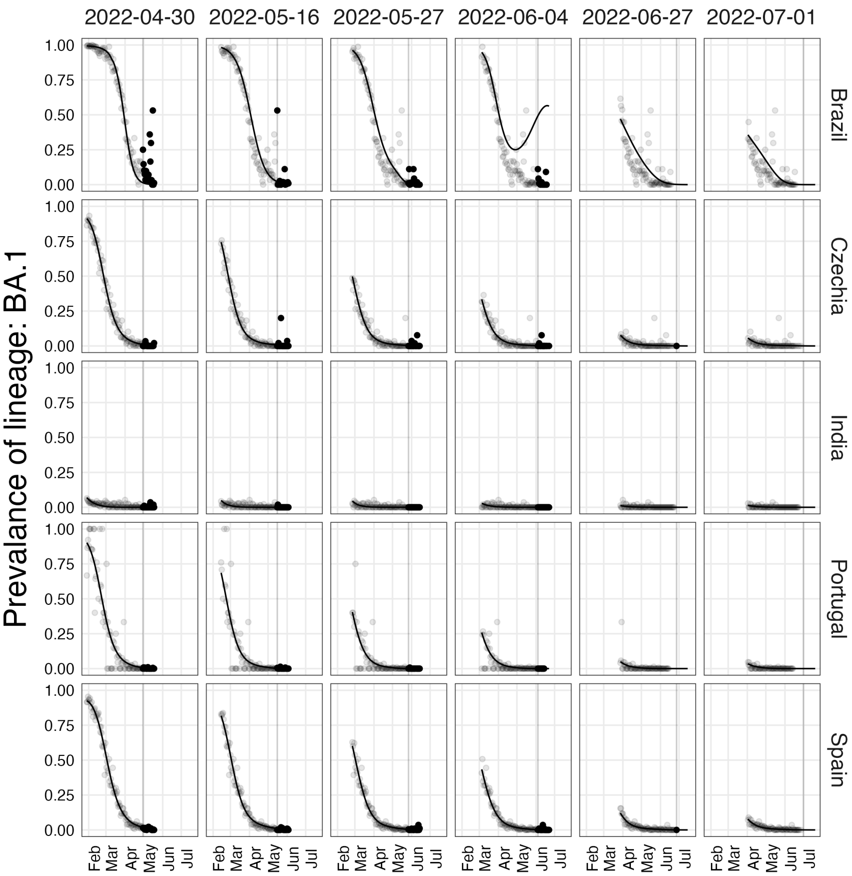

Figure S13: Predicted proportions of the BA.1 variant in countries of interest. The semi-transparent points are the "ground-truth" observed proportions in the dataset associated with the final reference date (not the reference date the model was fit on). The solid points are the 'ground-truth" observed proportions in the dataset associated with the final reference date in the 20 days after the last day with data that the model is fit to. The grey vertical bar is the last day with data that the model is fit to – the reference date. The black line are the predicted values in the country from the spline model fit to the reference date's dataset.

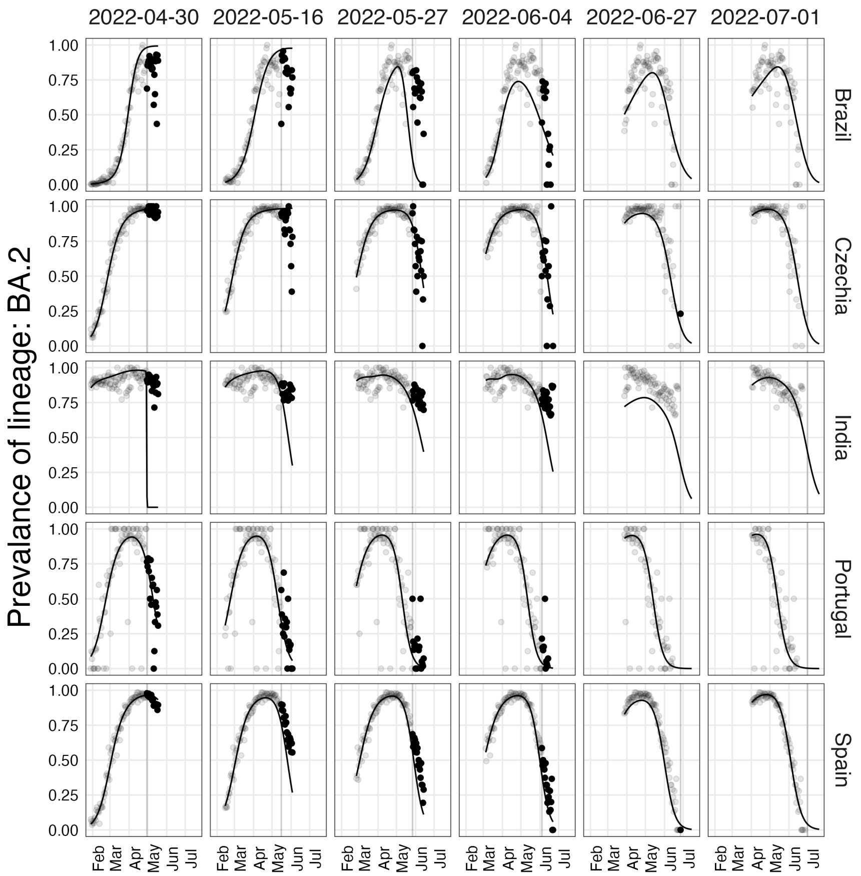

Figure S14: Predicted proportions of the BA.2 variant in countries of interest. The semi-transparent points are the "ground-truth" observed proportions in the dataset associated with the final reference date (not the reference date the model was fit on). The solid points are the 'ground-truth" observed proportions in the dataset associated with the final reference date in the 20 days after the last day with data that the model is fit to. The grey vertical bar is the last day with data that the model is fit to – the reference date. The black line are the predicted values in the country from the spline model fit to the reference date's dataset.

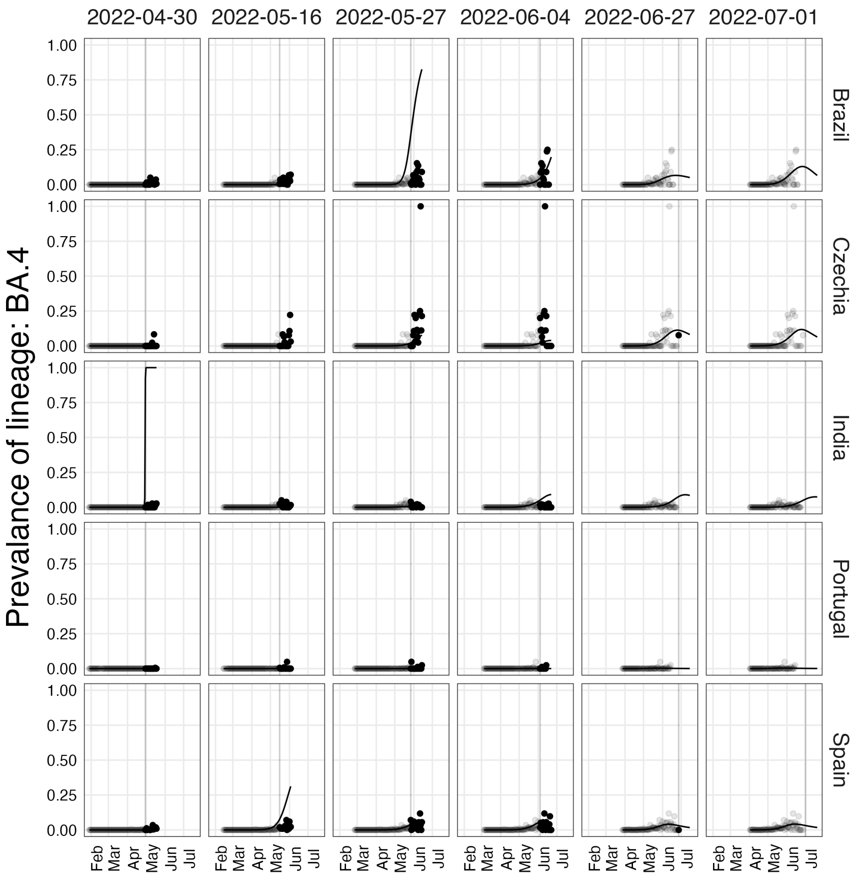

Figure S15: Predicted proportions of the BA.4 variant in countries of interest. The semi-transparent points are the "ground-truth" observed proportions in the dataset associated with the final reference date (not the reference date the model was fit on). The solid points are the 'ground-truth" observed proportions in the dataset associated with the final reference date in the 20 days after the last day with data that the model is fit to. The grey vertical bar is the last day with data that the model is fit to – the reference date. The black line are the predicted values in the country from the spline model fit to the reference date's dataset.

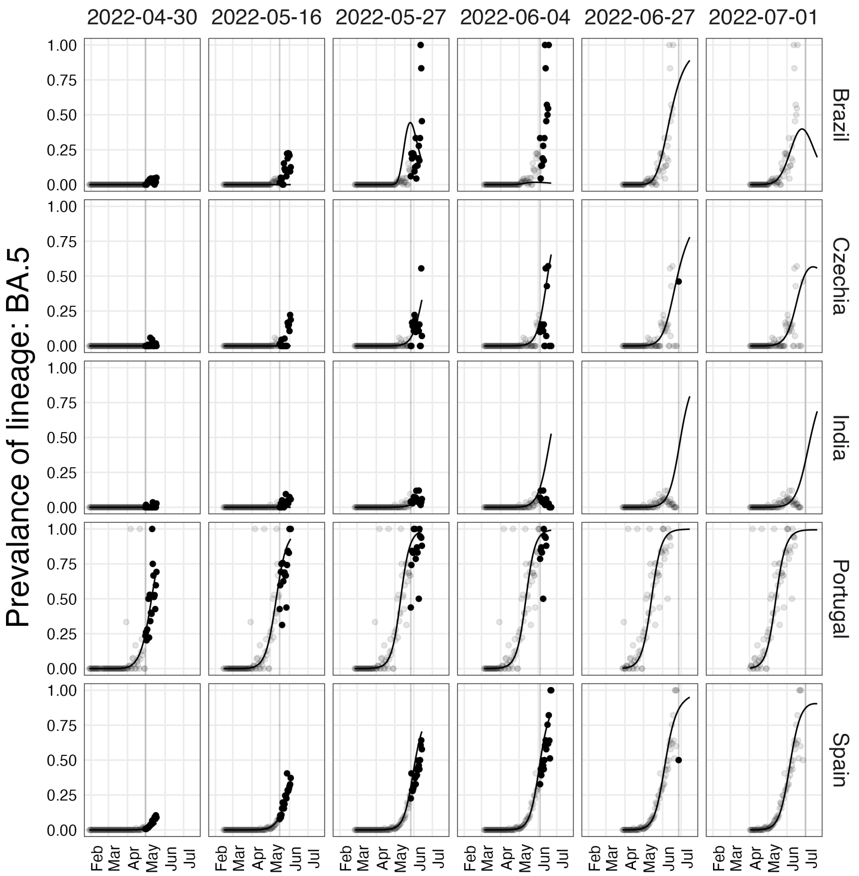

Figure S16: Predicted proportions of the BA.5 variant in countries of interest. The semi-transparent points are the "ground-truth" observed proportions in the dataset associated with the final reference date (not the reference date the model was fit on). The solid points are the 'ground-truth" observed proportions in the dataset associated with the final reference date in the 20 days after the last day with data that the model is fit to. The grey vertical bar is the last day with data that the model is fit to – the reference date. The black line are the predicted values in the country from the spline model fit to the reference date's dataset.

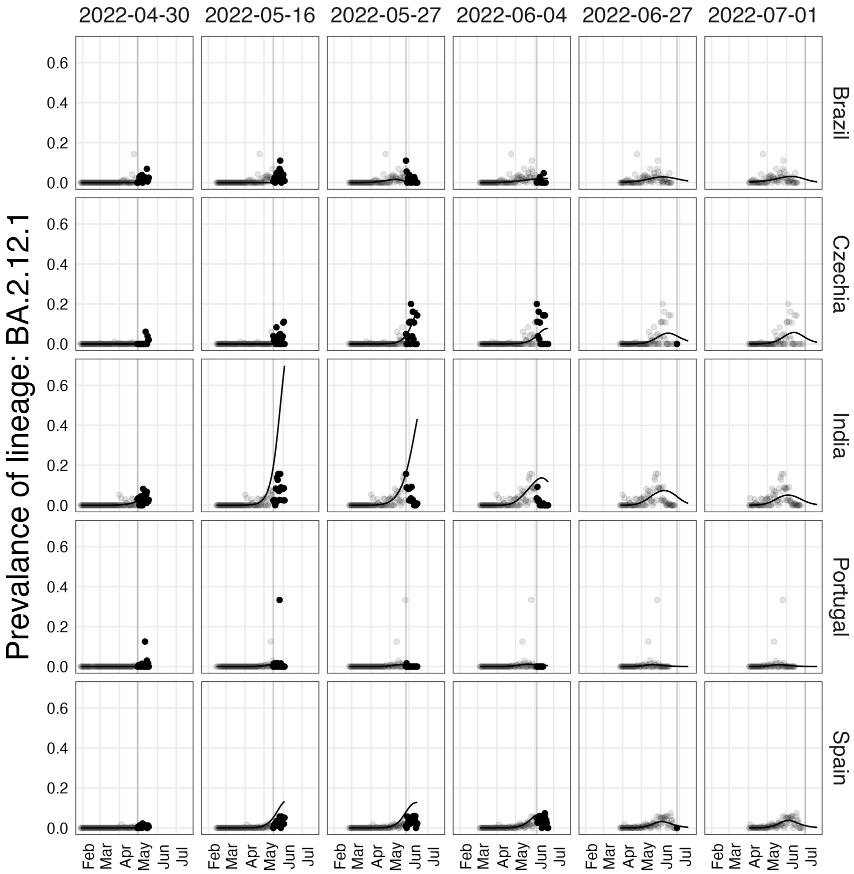

Figure S17: Predicted proportions of the BA.2.12.1 variant in countries of interest. The semi-transparent points are the "ground-truth" observed proportions in the dataset associated with the final reference date (not the reference date the model was fit on). The solid points are the 'ground-truth" observed proportions in the dataset associated with the final reference date in the 20 days after the last day with data that the model is fit to. The grey vertical bar is the last day with data that the model is fit to – the reference date. The black line are the predicted values in the country from the spline model fit to the reference date's dataset.

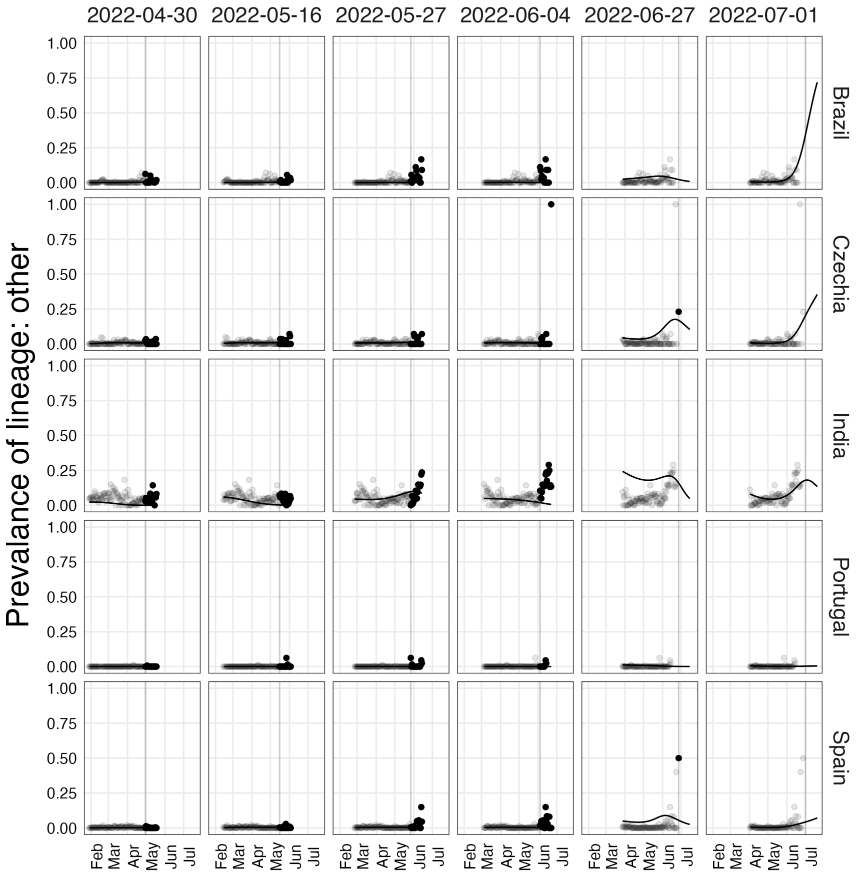

Figure S18: Predicted proportions of all other variants in countries of interest. These are all sequences that are not BA.1, BA.2, BA.12.12.1, BA.4 or BA.5. These predictions are computed by combining the predictions for the ”other” category in the model and the predictions for variants specified in the model but not one of the listed variants of interest. The semi-transparent points are the “ground-truth” observed proportions in the dataset associated with the final reference date (not the reference date the model was fit on). The solid points are the ‘ground-truth” observed proportions in the dataset associated with the final reference date in the 20 days after the last day with data that the model is fit to. The grey vertical bar is the last day with data that the model is fit to – the reference date. The black line are the predicted values in the country from the spline model fit to the reference date's dataset.

### 3 Case studies

#### 3.1 Identifying hemispheric drivers of influenza dynamics

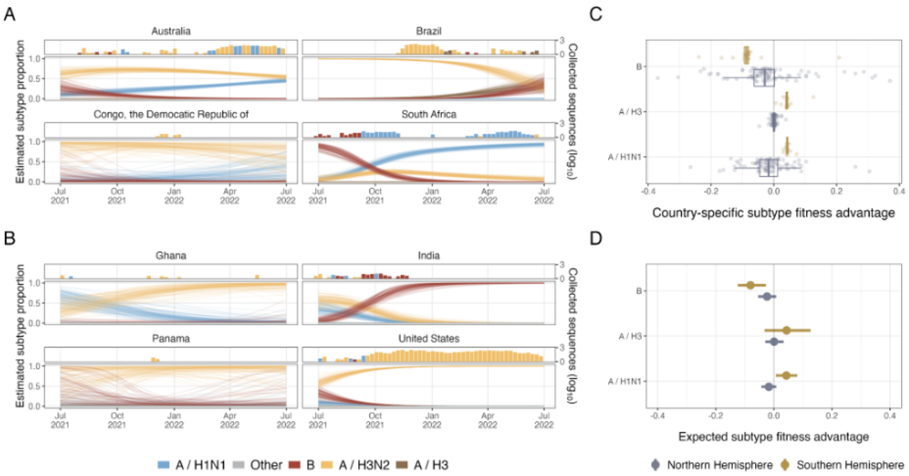

Figure S19: Application of the framework to Influenza dynamics in both hemispheres. (A) Flu variant dynamics for a subset of Southern Hemisphere countries. (B) Flu variant dynamics for a subset of Northern Hemisphere countries. (C) Country-specific fitness advantages relative to A / H3N2 for selected subtypes. (D) Expected fitness advantages relative to A / H3N2

We apply this approach to influenza dynamics, estimating fitness and subtype dynamics separately for the Northern and Southern hemispheres. In Figure S19A, we show estimates of influenza subtype prevalence from July, 2021 to July, 2022, identifying diverse dynamics across the selected countries from the Southern Hemisphere. In Australia and South Africa, prevalence of the B subtype steadily declines over the observed year and is replaced by a combination of A / H1N1 and A / H3N2. These dynamics differ from those of Brazil, where the dominant A / H3N2 subtype is replaced by A / H3 and B subtypes. Dynamics in the DRC are estimated imprecisely, but suggest that the A / H3N2 is dominant on July 1, 2022 — matching overall influenza dynamics in the Southern Hemisphere and the data observed from within the DRC.

In Figure S19B, we present estimated influenza subtype dynamics in the Northern Hemisphere from July 1, 2021 to July 2022. We find the A / H3N2 subtype increased in prevalence in the United States, Ghana, and Panama over the observed year. In all the selected countries, A / H1N1 prevalence declined or remained negligible. Notably, however, India's dynamics were meaningfully different from those of the other selected countries: the relative proportion of the B subtype increased from July, 2021 to July, 2022. It displaced both A /

H1N1 and A / H3N2, unlike in the other selected countries. We note that in many of these cases, influenza dynamics in the Northern Hemisphere appear to lag those of the Southern Hemisphere. In much of the Southern Hemisphere, the A / H3N2 and B subtypes are initially dominant and replaced over the observed year. In the Northern Hemisphere, the A / H3N2 and B subtypes are a small proportion of initial observations and increase in relative proportion throughout the observed year.

In Figure S19C and Figure S19D, we present estimates of country-specific and overall hemispheric mean fitness advantages for selected influenza subtypes. Because separate models are fit to the Northern and Southern Hemispheres, we present estimates for subtype fitness advantage relative to A / H3N2 separately for each hemisphere. We find that estimates of country-specific fitness advantage for the A / H3N2 are more diffuse in the Northern Hemisphere than in the Southern Hemisphere for all 3 subtypes. Except one outlier, estimated fitness advantages for all lineages are tightly clustered in the Southern Hemisphere. The Northern Hemisphere's more diffuse estimates have epidemiological implications: the estimates are scattered on either side of 0, indicating that the variance in country-specific fitness relative to A / H3N2 is high and any naive estimate for an unobserved country will be highly uncertain. The mean fitness advantages are similar for the B, A / H3, and A / H1N1 subtypes across hemispheres. For all three, estimates are either so close that the difference of means is statistically indistinguishable or close enough to not be epidemiologically meaningful. These results indicate that the country-specific differences are more important to influenza dynamics than hemisphere-level differences in subtype fitness.

#### 3.2 Estimating SARS-CoV-2 dynamics at administration level 1

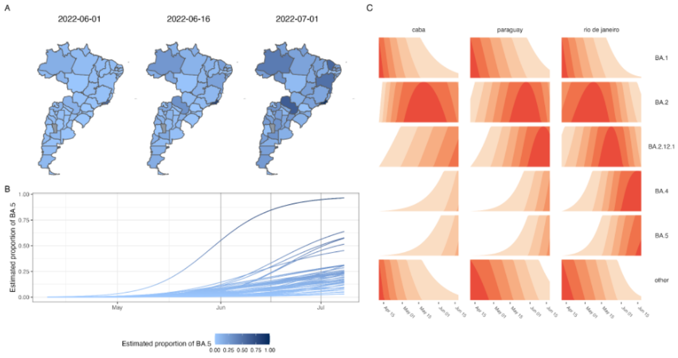

Figure S20: Application of the multicountry model to sub-country administrative units in Brazil, Paraguay, and Argentina

We apply the method described here to estimate SARS-CoV-2 dynamics at the AL0 and AL1 methods to demonstrate the flexibility of this modeling approach. We aggregate SARS-CoV-2 sequencing counts in GISAID from the states of Brazil and the provinces of Argentina (i.e., AL1 units) as well as combining sequences from all of Paraguay. These selections demonstrate how one might apply this method to estimate sub-national dynamics, where such data exist, and illustrate that the modeling approach remains coherent when estimating AL1 and AL0 spatial units together.

In Figure Figure S20A and Figure S20B, we show SARS-CoV-2 dynamics in these spatial units across time. We find that the BA.5 variant was uncommon across all the selected spatial units on June 6, 2022. Estimated variant prevalence was below 1% in all the modeled spatial units (Figure S20A). The expected prevalence of BA.5 rose over the course of the time period, increasing steadily through mid-June and into July (Figure S20B). We found, however, marked heterogeneity in the expected prevalence of BA.5 across the spatial units. BA.5 increased most quickly in Rio de Janeiro, approaching fixation by July 1, 2022. Although BA.5 prevalence was expected to have been increasing in all spatial units, BA.5 prevalence was below 50% on July 1, 2022 in most other regions in Brazil and Argentine.

In Figure S20C, we show prevalence over the observed time period for selected variants in three selected spatial units (CABA: Buenos Aires, Paraguay, and Rio de Janeiro). We identify the decline of BA.1 in all three of the spatial units during the time period. We find that BA.2 and BA.2.12.1 peaked in late May and early June and that it beginning to be outcompeted by BA.4 and BA.5. We also find that BA.4 had an earlier foothold in the region than BA.5.

This example demonstrates the ability of this applied method to be applied across heterogeneous spatial units. It identifies dynamics of SARS-CoV-2 variants and allows for variation across the spatial units in its estimates.

### 4 Data/code availability

All code is made publicly available at this Github repository Github repository. We do not include any data in the repository, but all results can be reproduced using the GISAID SARS-CoV-2 and flu metadata for authenticated users.

### 5 EPI_SET identifier

**Table S1: GISAID EPI_SET identifier that uniquely identifies all sequences used in this manuscript**. All sequencing labs are credited along with the associated SARS-CoV-2 sequences at the link associated with the DOI.

### SUPPLEMENTAL TABLE

**Data Availability**

| | |
|---|---|
| GISAID Identifier: | EPI_SET_230118ka |
| doi: | 10.55876/gis8.230118ka |

All genome sequences and associated metadata in this dataset are published in GISAID's EpiCoV database. To view the contributors of each individual sequence with details such as accession number, Virus name, Collection date, Originating Lab and Submitting Lab and the list of Authors, visit 10.55876/gis8.230118ka

**Data Snapshot**

- EPI_SET_230118ka is composed of 2,032,779 individual genome sequences.
- The collection dates range from 2022-04-01 to 2022-08-01;
- Data were collected in 168 countries and territories;
- All sequences in this dataset are compared relative to hCoV-19/Wuhan/WIV04/2019 (WIV04), the official reference sequence employed by GISAID (EPI_ISL_402124). Learn more at https://gisaid.org/WIV04.

### References

[1] *cmdstan: CmdStan, the command line interface to Stan*. en.

[2] *COG-UK*. https://pangolin.cog-uk.io/. Accessed: 2022-9-15.

[3] Nicholas G Davies et al. "Estimated transmissibility and impact of SARS-CoV-2 lineage B.1.1.7 in England". en. In: *Science* 372.6538 (Apr. 2021).

[4] Marlin D Figgins and Trevor Bedford. "SARS-CoV-2 variant dynamics across US states show consistent differences in effective reproduction numbers". Dec. 2021.

[5] Shruti Khare et al. "GISAID's role in pandemic response". In: *China CDC Weekly* 3.49 (2021), p. 1049.

[6] Russell V. Lenth. *emmeans: Estimated Marginal Means, aka Least-Squares Means*. R package version 1.8.5. 2023. URL: https://CRAN.R-project.org/package=emmeans.

[7] Fritz Obermeyer et al. "Analysis of 6.4 million SARS-CoV-2 genomes identifies mutations associated with fitness". In: *Science* 376.6599 (2022), pp. 1327–1332.

[8] *pangoLEARN*. en.

[9] Sang Woo Park et al. "The importance of the generation interval in investigating dynamics and control of new SARS-CoV-2 variants". In: *Journal of The Royal Society Interface* 19.191 (2022), p. 20220173.

[10] Prabasaj Paul et al. "Genomic Surveillance for SARS-CoV-2 Variants Circulating in the United States, December 2020-May 2021". en. In: *MMWR Morb. Mortal. Wkly. Rep.* 70.23 (June 2021), pp. 846–850.

[11] Brian Ripley. "Feed-Forward Neural Networks and Multinomial Log-Linear Models [R package nnet version 7.3-17]". In: (Jan. 2022).

[12] *stan Wiki*. en.

[13] Bradford P Taylor and William P Hanage. "A simple model of how heterogeneous disease transmission impacts the emergence of variants and their detection". Nov. 2022.

[14] *Tracking SARS-CoV-2 variants*. https://www.who.int/activities/tracking-SARS-CoV-2-variants. Accessed: 2023-3-10. Mar. 2022.

[15] Harald S Vöhringer et al. "Genomic reconstruction of the SARS-CoV-2 epidemic across England from September 2020 to May 2021". In: *medRxiv* (2021), pp. 2021–05.

[16] Tom Wenseleers. *Tomwenseleers/LineageExplorer: Estimate growth rate advantage of SARS-cov2 variants of concern based on International Genomic Surveillance Data and multinomial spline fits*. URL: https://github.com/tomwenseleers/LineageExplorer.

[17] Simon N Wood. *Generalized additive models: an introduction with R*. chapman and hall/CRC, 2006.
